# MIS-CYTO: A prospective, multi-center, observational study for validation of oncological adequacy of minimally invasive cytoreductive surgery for peritoneal malignancies with limited peritoneal spread at systematic mini-laparotomy by INDEPSO

**DOI:** 10.64898/2026.05.23.26353919

**Authors:** Mufaddal Kazi, Vivek Sukumar, Sanket Mehta, Ajinkya Pawar, Swapnil Patel, Deepti Mishra, Niharika Garach, Praveen Kammar, Zeeba Usofi, Snita Sinukumar, Rajagopalan R Iyer, Vivekanand Sharma, Ambarish Chatterjee, Ankita Patel, Rohit Ranade, Aruna Prabhu, Dileep Damodaran, AS Ramakrishnan, Shigeki Kusamura, Avanish Saklani, Kurt Van Der Speeten, Aditi Bhatt

**Affiliations:** Dept of Colorectal and Gastrointestinal Surgery, Tata Memorial Hospital, Mumbai, India; Dept of Surgical Oncology, Specialty Surgical Oncology, Mumbai, India; Dept of Surgical Oncology, Gujarat Cancer and Research Institute, Ahmedabad, India; Dept of Surgical Oncology, Upar Hospital and Cancer Institute, Varanasi, India; Dept of Surgical Oncology, Thangam cancer center, Nammakal, India; Dept of Surgical Oncology, Basvatarakam Indo-American Cancer Hospital, Hyderabad, India; Dept of Surgical Oncology, Apollo Hospital, Pune, India; Dept of Surgical Oncology, Fortis Hospital, Mumbai, India; Dept of Surgical Oncology, Apollo Hospital, Nashik, India; Dept of Surgical Oncology, Specialty Surgical Oncology, Ahmedabad, India; Dept of Gynecological Oncology, Narayana Health City, Bangalore, India; Dept of Surgical Oncology, MVR cancer center and research Institute, Kozhikode, India; Integrative Cancer Care Group, Chennai, India; Department of Surgical Oncology, Fondazione IRCCS Istituto Nazionale dei Tumori, Milano, Italy; Department of Surgical Oncology, Ziekenhuis Oost-Limberg, Genk, Belgium; University Hasselt BIOMED Research Institute Faculty of Life Sciences, Hasselt, Belgium; Dept of Surgical Oncology, Shalby Cancer and Research Institute, Ahmedabad, India

## Abstract

**Background:** The role of minimally-invasive cytoreductive surgery (MI-CRS) in low volume peritoneal disease has been explored but lacks robust evidence demonstrating oncological adequacy and non-inferiority to open CRS. The major challenges of MI-CRS are its technical complexity and chances of missing disease which could have detrimental oncological outcomes. The only means of resolving the dilemma of missed disease is to convert the procedure to open surgery and confirming the findings of MI approach. The “Minimally InvaSive CYTOreductive surgery” (MIS-CYTO) study is a prospective observational study that will evaluate the oncological adequacy of MI-CRS at systematic mini-laparotomy in patients with low volume peritoneal disease.

**Methods:** This single-arm study which will include 100 patients with primary and secondary peritoneal malignancies with a peritoneal cancer index (PCI)< 10 treated by MI-CRS. Patients will be recruited over 3 years and followed up for 5 years. All patients will undergo a staging laparoscopy (SL) with video documentation according to the study format. Patients who undergo subsequent MI-CRS will be included in the per-protocol population. At the end of MI-CRS video documentation of the resection sites is performed followed by a systematic mini-laparotomy for exploration of the abdominal cavity for any residual disease. The presence of residual disease will be confirmed on pathological evaluation. The primary end point is the incidence of missed disease during MI-CRS that is detected during mini-laparotomy. Quality assessment of SL and MI-CRS will be performed by an independent two-member committee. Patients undergoing conversion to open cytoreductive surgery will comprise the comparator group for studying the secondary end-point which include the sites of missed disease, perioperative outcomes and survival outcomes.

Survival endpoints will be measured from the date of surgery to the event: overall survival (OS) will be defined by death due to any cause, and progression-free or relapse-free survival (PFS/RFS) by any recurrence or death whichever comes first.

**Ethics and Registration:** The study protocol is approved by the ethics committee of Shalby Cancer and Research Institute on 7^th^ October 2024 (EC/069/02). The study is registered under the Clinical Trials Registry of India; subsidiary of clinicaltrials.gov (CTRI/2024/11/076312).

**Strengths and limitations:**

- The study will be the first to demonstrate the oncological adequacy or inadequacy of the MI-CRS
- The methodology and surgical protocol are robust and should minimize heterogeneity in surgical practices in the study
- If the oncological inadequacy of MI-CRS is not demonstrated, its use should be limited in patients with limited peritoneal disease.
- Two independent experts will review the adequacy of staging laparoscopy and MI-CRS both
- The stringent methodology could be a deterrent for surgeons/ centres to participate in the study, specifically performing a systematic mini-laparotomy for those surgeons who prefer to deliver specimens through a natural orifice.

## Introduction

Cytoreductive surgery (CRS), performed with the objective of achieving a complete cytoreduction (CC-0 resection), is the cornerstone of life-prolonging and/or potentially curative treatment for peritoneal malignancies. (1,2) Conventionally, CRS is performed via a xipho-pubic laparotomy. The morbidity of CRS is often higher compared to other oncological surgeries but varies greatly according to the extent of surgery that is performed. (3) Minimal access surgery has revolutionized the treatment of abdominal and thoracic malignancies, with faster recovery and oncological non-inferiority being clearly demonstrated for many procedures in well-designed clinical studies and randomized trials. (4–10) The role of minimally-invasive cytoreductive surgery (MI-CRS) in patients with limited or low-volume peritoneal disease has been explored. The early results are encouraging but robust evidence demonstrating shorter postoperative recovery times and oncological non-inferiority is lacking. (11,12) Most of the evidence comes from retrospective cohort studies with a limited number of patients. (13–15)

There are two major challenges in MI-CRS. First, performing peritonectomy procedures by the MIS approach is technically challenging and time-consuming. The techniques of most peritonectomy procedures have not been described in published literature. Clearance of disease from some regions like the superior recess of the lesser sac, Foramen-of-Winslow, and bowel mesentery may not be possible by this approach. (16) The robotic approach may reduce the technical challenges in performing some procedures. (17) The second and most critical challenge is assessing the disease extent and residual disease accurately. In contrast to the other minimally invasive surgeries, where the surgical extent is guided by preoperative imaging and oncological adequacy is subsequently confirmed through histopathological evaluation, the only metric to evaluate the quality of cytoreductive surgery is the surgeon’s documentation of the completeness of cytoreduction. This evaluation is entirely based on the intraoperative assessment of residual disease by visual inspection and palpation. Owing to the absence of tactile feedback in minimally invasive surgery and the limited visualization of certain areas, there exists risk of missing disease during the procedure. This may have detrimental consequences on patient outcomes, especially in cases of high-grade malignancies such as colorectal and ovarian cancers.

The only means of resolving the dilemma of missed disease and validating the feasibility and oncological adequacy of minimally invasive CRS is to convert the procedure to open surgery and confirming the findings of minimally invasive approach. (18) With this rationale in mind, the MIS-CYTO (MInimally invaSive CYTOreductive surgery) study was conceived. The main goal is to study the incidence of missed peritoneal disease when MI-CRS is performed in well selected patients. The secondary goals are to study the rate of conversion to laparotomy to achieve a complete cytoreduction and the reasons for conversion, the sites where disease was missed during MI-CRS, which peritonectomies/resections can be performed during MI-CRS and the perioperative and survival outcomes after MI-CRS.

## Methods

This study is a single-arm, prospective, multi-centre observational study designed to evaluate the oncological adequacy of minimally invasive cytoreductive surgery (MI-CRS) in patients with peritoneal malignancies by performing a systematic mini-laparotomy at the end of MI-CRS. The study will be carried out at 10 Indian peritoneal malignancy centers. Participation of centres outside India is permitted. Patients with both primary and secondary peritoneal cancers will be eligible for inclusion if they meet the following criteria: a peritoneal cancer index (PCI) of less than 10 and no radiological evidence of unresectability. However, patients with a low PCI (<10) in whom a complete cytoreduction (CC-0/1) cannot be achieved will be excluded from the study. Additionally, individuals with a low PCI on imaging in whom MI-CRS is deemed unfeasible such as those with a history of prior CRS via laparotomy, multiple extensive laparotomies, or contraindications to a minimally invasive approach will not be enrolled in the study **(Figure 1).** Baseline data of these patients will still be collected to the proportion of individuals with low PCI who are ineligible for MI-CRS. All patients scheduled for MI-CRS will first undergo a staging laparoscopy (SL), either immediately preceding the MI-CRS or as a separate intervention, when clinically permissible. The SL findings will be systematically documented in a pre-specified format detailing the condition of each region of the peritoneal cavity **(Table 1)**. These recordings will be submitted to the regulatory committee for evaluation. Each of the two committee members will independently assess the quality of the video documentation. Patients with evidence of unresectable disease identified during laparoscopy will be excluded from the study; however, baseline data will still be collected for these cases. The study imposes no restrictions on the sites of disease eligible for resection, provided the operating surgeon is confident in performing the procedure via the minimally invasive approach. MI-CRS may be conducted using either laparoscopic or robotic techniques. Upon completion of the cytoreductive surgery, a video recording of the resection sites, demonstrating the completeness of the cytoreduction, will be submitted to the regulatory committee. Each of the two committee members will independently review the quality of this video documentation **(Table 2)**. Following MI-CRS, in cases where a CC-0/1 resection is achieved, the abdominal cavity will be explored via a midline mini-laparotomy. The incision will extend from a midpoint between the xiphoid process and the umbilicus to a midpoint between the umbilicus and the pubic symphysis. All resected specimens will be sent for pathological evaluation. In cases where hyperthermic intraperitoneal chemotherapy (HIPEC) is indicated, it will be administered following the open portion of the procedure, using either the open or closed technique, according to the surgeon’s standard practice. If the surgeon determines that conversion to laparotomy is necessary to achieve complete cytoreduction, the procedure will be converted accordingly. Patients requiring conversion to laparotomy prior to achieving a CC-0/1 resection will be excluded from the study; however, comprehensive data, including the reason for conversion, will be documented. All additional procedures performed in patients who require conversion to laparotomy prior to a CC-0/1 resection will be meticulously recorded. Post-operative outcomes will be recorded and include the intensive care unit-stay, hospital stay, grade 3-4 complications at 30 and 90-days, grade 1-2 complications, 90-day mortality, time-to-return of bowel function. The perioperative pain management will be as per the existing institutional protocol but will be captured in detail. The study design is illustrated in **Figure 1 and protocol provided as a supplementary file**.

**Figure 1:**
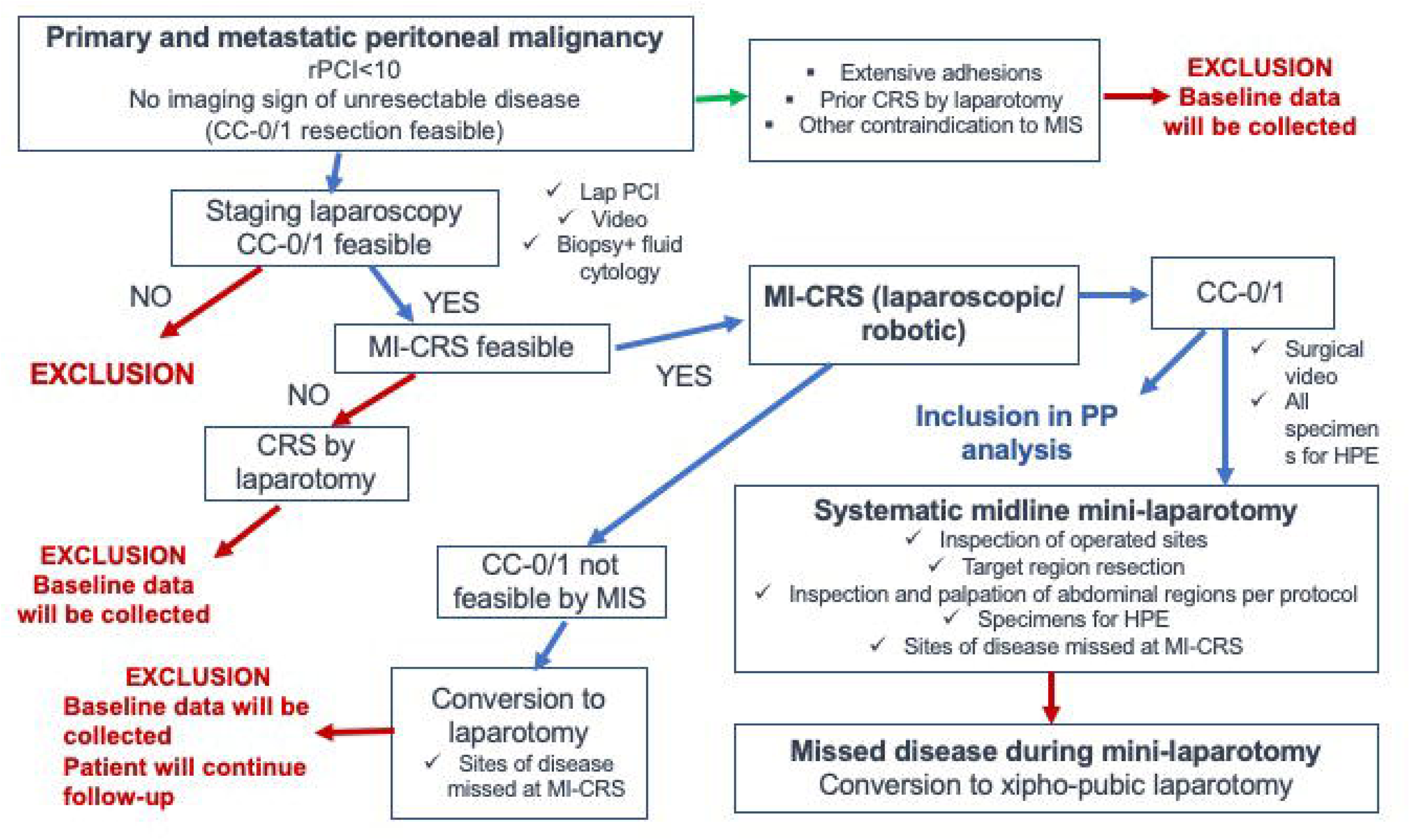
Design of the MIS-CYTO study Abbreviations: PP-per protocol

**Table 1:**
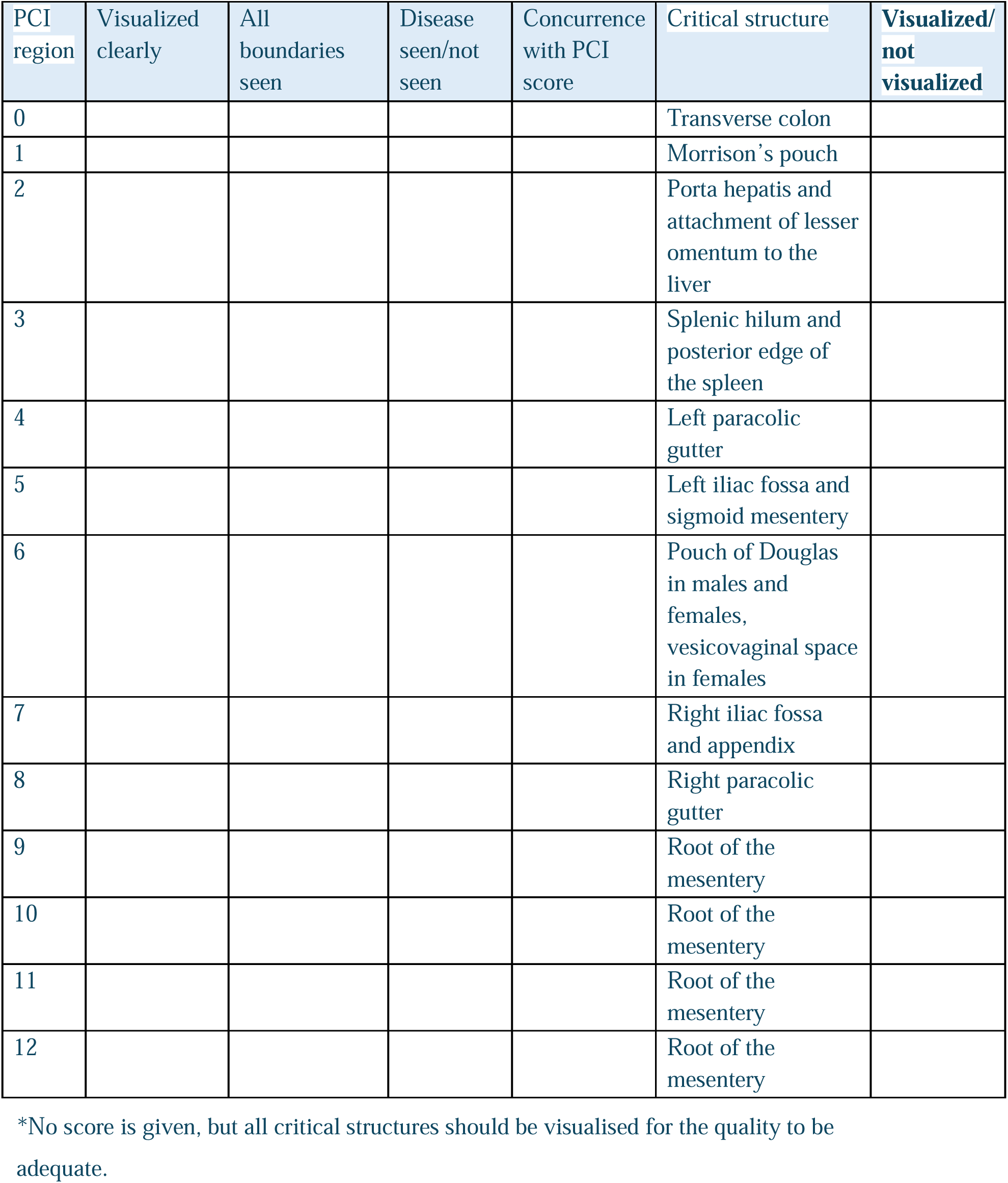
Format for assessing the quality of the staging laparoscopy videos*.

**Table 2:**
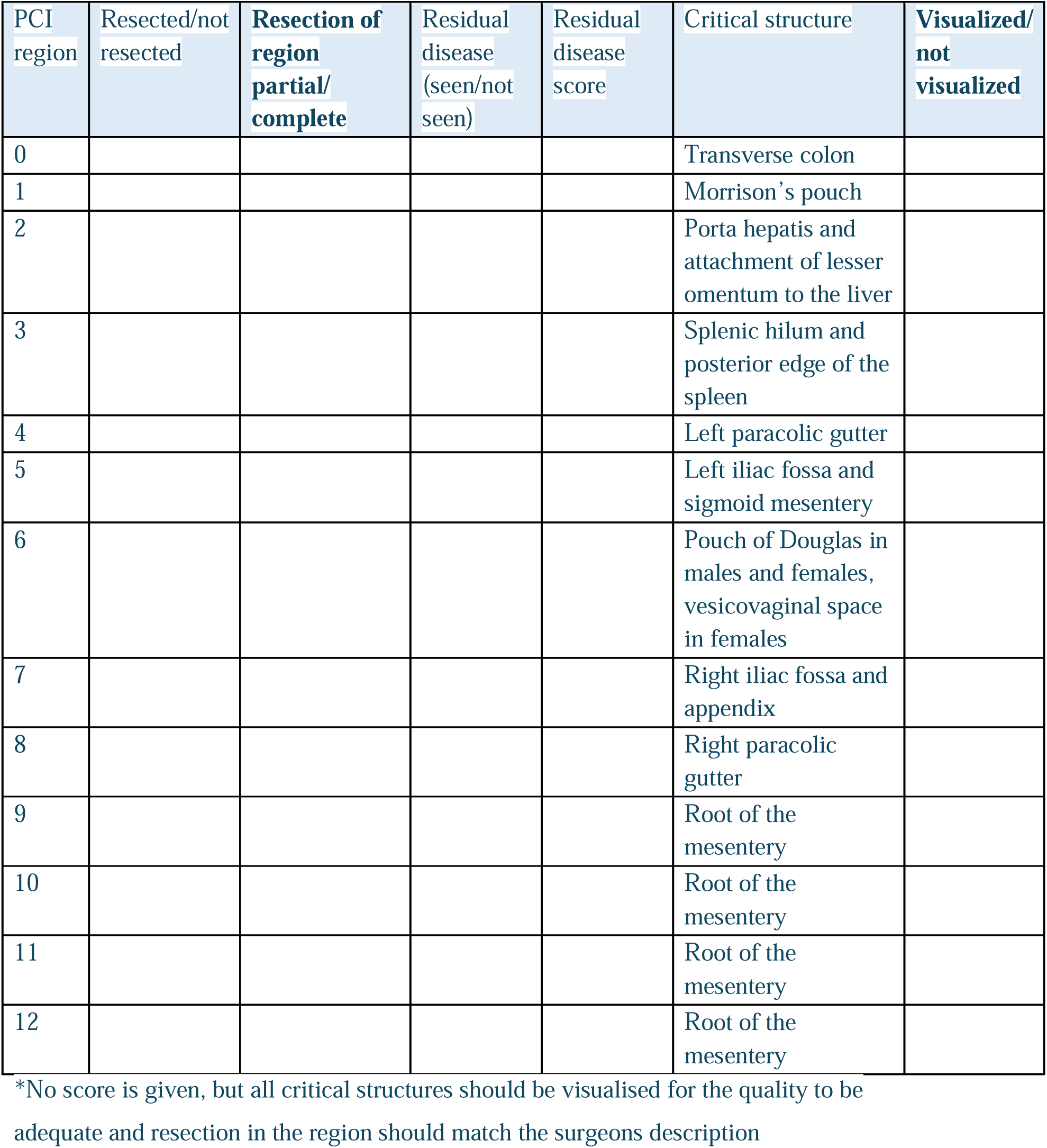
Format for assessing the quality of the MI-CRS videos*.

| PCI region | Resected/not resected | Resection of region partial/complete | Residual disease (seen/not seen) | Residual disease score | Critical structure | Visualized/not visualized |
| --- | --- | --- | --- | --- | --- | --- |
| 0 |  |  |  |  | Transverse colon |  |
| 1 |  |  |  |  | Morrison's pouch |  |
| 2 |  |  |  |  | Porta hepatis and attachment of lesser omentum to the liver |  |
| 3 |  |  |  |  | Splenic hilum and posterior edge of the spleen |  |
| 4 |  |  |  |  | Left paracolic gutter |  |
| 5 |  |  |  |  | Left iliac fossa and sigmoid mesentery |  |
| 6 |  |  |  |  | Pouch of Douglas in males and females, vesicovaginal space in females |  |
| 7 |  |  |  |  | Right iliac fossa and appendix |  |
| 8 |  |  |  |  | Right paracolic gutter |  |
| 9 |  |  |  |  | Root of the mesentery |  |
| 10 |  |  |  |  | Root of the mesentery |  |
| 11 |  |  |  |  | Root of the mesentery |  |
| 12 |  |  |  |  | Root of the mesentery |  |

\*No score is given, but all critical structures should be visualised for the quality to be adequate and resection in the region should match the surgeons description

## Inclusion criteria

- Patients with biopsy proven peritoneal malignancy from the following primary sites: colorectal cancer, peritoneal mesothelioma, pseudomyxoma peritonei (PMP) and gastric cancer
- Patients with advanced epithelial ovarian cancer undergoing primary or interval cytoreductive surgery
- Radiological PCI < 10 with no sign of unresectability on imaging
- Laparoscopic PCI < 10
- Recurrent ovarian cancer undergoing secondary cytoreductive surgery with disease at 1-2 sites only.
- Patients who have had previous laparoscopic surgery for primary tumor resection will be included if all the other criteria are fulfilled
- Patients with prior laparotomy could be included if the same was not performed for treating peritoneal malignancy by cytoreductive surgery
- Age 18-70 years
- Normal blood counts and blood biochemistry
- No medical contraindication to major abdominal surgery

## Exclusion criteria

- Prior CRS+/-HIPEC
- Patients with a PCI<10 with unresectable disease on imaging or at staging laparoscopy
- Patients undergoing prophylactic procedures/second look procedures
- Primary tumor site excluding colorectal cancer, gastric cancer, advanced epithelial ovarian cancer, multi-cystic peritoneal mesothelioma and PMP
- Patient refusing consent

## Endpoints

### Primary endpoint

The primary endpoint of this study is the proportion of patients with pathologically proven limited peritoneal disease that was not identified/resected during MI-CRS.

### Secondary endpoints

The secondary endpoints of this study are

- Incidence of pathologically proven disease in ‘normal appearing’ target regions (greater omentum, lesser omentum, falciform ligament and/or umbilical round ligament) resected during laparotomy which were not resected during MI-CRS
- Duration of hospitalization
- Grade 3-4 morbidity at 90 days
- Perioperative outcomes: operative time
- Perioperative outcomes: blood loss
- Perioperative outcomes: use of opioids
- Recovery of bowel function (time to first flatus)
- 1-year progression-free survival (PFS)
- 3-year PFS and overall survival (OS)
- 5-year PFS and OS
- Rate of conversion to laparotomy

### Study duration

The total study duration is 8 years. Recruitment should take 3 years and follow-up will continue for 5 years after recruitment of the last patient.

### Imaging protocol

CT scan with intravenous and/or oral contrast, FDG PET scan and/or peritoneal MRI are the imaging modalities for assessing the extent of peritoneal disease before surgery. (19,20) The combination of above may be performed when indicated. Where facilities and expertise are present, peritoneal MRI will be preferred modality. Both T1 and T2, gadolinium contrast enhanced and diffusion weighted images will be used to map the disease. (21) A radiological PCI will be calculated and patients screened for radiological signs of unresectable disease. (22) Preoperative imaging will be performed, preferably within 2-4 weeks of planned surgery. All scans performed will be considered while evaluating the patient.

### Surgical procedures

#### Staging laparoscopy

It should be performed as an independent procedure or just prior to commencing with MI-CRS. All ports or the procedure should be placed in midline whenever feasible. All 13 regions should be evaluated thoroughly and PCI calculated. The PCI will be reported according to the description of morphology of lesion provided in the form **(Supplement 1)** A video showing all the 13 regions of abdomen explored and assessed in the laparoscopy should be shot and submitted with the clinical data to the regulatory committee. This should also include areas of suspicion and uninvolved regions. Each of the two committee members will independently assess the quality of staging laparoscopy **(Table 1)**. Peritoneal biopsies should be taken to confirm malignancy if not done previously. Ascitic fluid if any will be sampled and in absence of ascites peritoneal washings after instillation of 200 ml of normal saline will be taken and sent for cytology and/or cell block analysis.

#### Minimally invasive cytoreductive surgery

This procedure could be performed laparoscopically or robotically. The port placement will be at the discretion of the surgeon. A staging laparoscopy will be performed as described above and findings recorded. If it was performed earlier, the findings would be confirmed. Cytoreductive surgery will then be performed to achieve a CC-0 resection. The sequence of procedures, technique of peritonectomy, visceral resections, use of frozen section, etc. will be according to the surgeon’s preference. The use of mini-laparotomy or any large incisions to palpate the disease will not be permissible at this stage of surgery as the main purpose of this study is to find the incidence of missed disease. After the completion of this phase a video showing all the sites of resection in full extent should be shot and submitted to the regulatory committee. Each of the two committee members is independently assess the quality of the MI-CRS (Table 2). The specimens will be delivered in the mini-laparotomy phase.

#### Mini-laparotomy phase

In this phase, the abdomen will be opened though a midline mini-laparotomy that extends from mid-way between the xiphoid process to the umbilicus to mid-way between the umbilicus and pubic symphysis. All the resected areas will be systematically inspected for the presence of any residual disease. The 13-region residual PCI will be recorded in the PCI form that has provision for the same. The whole colon and rectum will be palpated for the presence of nodules. The entire small bowel will be inspected for presence of tumor nodules. The lesser omentum will be inspected and palpated for presence of disease. If a splenectomy is not performed, then the splenic hilum and posterior edge will be both inspected for presence of disease. The Morrison’s pouch, gall bladder and foramen of Winslow will be inspected for presence of disease. Liver mobilization could be done if deemed necessary by the surgeon but is not mandatory. The ovaries if present will be inspected for presence of disease. A total omentectomy preserving the gastroepiploic arch will be performed for all patients even if absence of gross disease if the same was not done during MI-CRS. If the arch is directly involved or resection deemed necessary by the surgeon, it will be resected. The falciform ligament and umbilical round ligament will be resected in all patients irrespective of the presence of disease if the same was not done during MI-CRS. Any newly identified areas of disease will be resected performing the necessary peritonectomies and visceral resections. Conversion to full laparotomy should be done if necessary to resect the newly detected disease. The specimens will be sent for histopathological evaluation. All port sites of previous staging laparoscopy will be resected at the end of the procedure and sent for pathologic evaluation. At the end of the procedure, HIPEC will be performed if indicated by the open or the closed method according to the preference of the surgeon. The postoperative management will be according to the protocol at each institution.

### Pathological evaluation

The pathological specimen of each peritoneal region will be submitted separately and all regions will be individually analysed. Two or more representative should be taken from each region. Similarly, all the visceral specimens will be evaluated separately. (23) A pathological PCI will be calculated based on lesion size score of each region. This will be confirmed on microscopy. (24) A pathological response grade will also be mentioned if patient received any previous treatment prior to surgery. (25) Ascitic fluid or peritoneal washings will undergo cytologic analysis and cell block analysis if required. The reporting of pathological findings of primary tumour if present will be according to existing protocols of the centre.

### Additional treatment(s)

Additional systemic treatments will be at the discretion of the treating clinicians but will be captured and submitted for each case.

### Follow up/ surveillance

Routine follow-up of all patients will be done. Depending on the primary tumor site, surveillance will involve physical examination, blood tests and cross-sectional imaging as deemed necessary by the treating clinician. The frequency will be every 3-6 months for the first two years and every 6-12 months for the next three years, depending on the primary tumor site.

### Sample size

The essence of this study lies in detecting peritoneal metastases missed during MI-CRS. Given that the eligible patients will have low-volume, potentially curable disease, the margin for error must be minimal. Incomplete cytoreduction is the most significant negative prognostic factor in CRS, regardless of the primary tumour. Therefore, the incidence of missed metastases should ideally be 0%. Assuming a tolerable limit of ≤10% missed metastases, with an acceptable margin of ≤5%, a sample size of 88 patients is required to achieve 95% confidence. This ensures that if fewer than 4 patients have missed metastases, there is 95% certainty that the true incidence does not exceed 10%. To accommodate potential conversions during MI-CRS and unforeseen complications, a total of 100 patients will be recruited.

## Statistical methods

In open surgery, the incidence of missed metastases is considered to be zero. Therefore, any occurrence of missed disease in this study is deemed unacceptable. The primary endpoint does not require a comparative group. For the secondary endpoints, the comparator group will comprise patients excluded due to CRS contraindications, extensive adhesions, prior CRS by laparotomy, or unresectable disease identified during staging laparoscopy. Both primary and secondary endpoints will be analysed in the per-protocol population **(Table 3)**. Continuous variables will be reported as medians with interquartile ranges, while categorical variables will be presented as numbers and proportions. Survival endpoints will be measured from the date of surgery to the event: overall survival (OS) will be defined by death due to any cause, and progression-free or relapse-free survival (PFS/RFS) by any recurrence or death whichever comes first. Patients without events will be censored at their last follow-up. Time-to-event data will be analysed using the Kaplan-Meier method, while follow-up durations will be estimated using the reverse Kaplan-Meier method.

**Table 3.** Schema for evaluation of primary and secondary end-points.

| Outcomes | Timing | Population |
| --- | --- | --- |
| **Primary outcome** |  |  |
| Disease missed at MI-CRS | Yearly | Per protocol |
| **Secondary outcomes** |  |  |
| Occult disease in target regions at laparotomy | Yearly | Per protocol |
| Hospital stay | Yearly | Per protocol |
| Grade 3-4 complications | Yearly | Per protocol |
| Perioperative outcomes- operative time, blood loss, time to first flatus and opioid use | Yearly | Per protocol |
| 1- year progression-free survival (PFS) | 1 year post enrollment | Per protocol |
| 3- year PFS and overall survival (OS) | 3 years post enrollment | Per protocol |
| 5- year PFS and OS | 5 years post enrollment | Per protocol |
| Rate of conversion of laparotomy | Yearly | Per protocol |
| Incidence of pathologically proven disease in ‘normal appearing’ target regions resected during laparotomy | Yearly | Per protocol |
| Site of residual disease during mini-laparotomy | Yearly | Per protocol |
| Peritonectomies and visceral resections performed by MIS and post resection video acceptable by the regulatory committee | Yearly | Per protocol |

## Ethics and dissemination

This study is sponsored by the Indian Network for Development of Peritoneal Surface Oncology (INDEPSO) and approved by the ethics committee of Shalby Cancer and Research Institute on 7^th^ October 2024 (EC/069/02), the institution of the first author and principle investigator. The study is registered under the trial registry number CTRI/2024/11/076312 (Clinical Trials Registry of India; subsidiary of clinicaltrials.gov) and has been recruiting patients since 1^st^ December, 2024.

At the end of follow up phase, proposed analysis will be carried out and the results will be published in peer reviewed journal.

### Patient and public involvement

A written informed consent will be obtained from all patients for their enrolment in this study. Patients must be able to understand and comply with written and verbal protocol requirements, instructions, and protocol-stated restrictions. Patients should also be able to sign and date the written informed consent document acknowledging their desire to participate in the study, or if unable to sign, a legal representative consent must be obtained before the stipulated intervention. The results of the study will be published in appropriate scientific journals upon completion of the study. The results will be disseminated on social media platforms by the administrative body INDEPSO (Indian Network for Development of Peritoneal Surface Oncology) to disseminate the results among clinicians and laypersons alike.

The SPIRIT checklist is provided as Supplement 1

## Discussion

There has been a paradigm shift from open surgery to minimally invasive surgery (MIS) for the management of a wide range of malignancies, supported by robust evidence demonstrating its oncological adequacy and non-inferiority. With an increase in the awareness of peritoneal metastases and the advent of newer imaging techniques and protocols, peritoneal malignancy is detected early in many patients with limited spread and low volumes. (2) The application of minimally invasive CRS for this population of patients has been explored and seems alluring. Apart from better perioperative short term outcomes, other advantages like lesser ‘fibrin entrapment’, decreased over all costs, higher pressures increasing penetrance and cytotoxicity of HIPEC and decreased ‘time to adjuvant chemotherapy’ have been proposed which may potentially influence the long term outcomes. (12, 26–29) However, there is no robust evidence to back this proposition as majority of evidence comes from small retrospective studies. There are two randomized trials comparing minimally invasive and open CRS, the LANCE trial and the Mirrors trial for interval CRS in advanced ovarian cancer. (30,31) In the LANCE trial only patients who have a complete or near complete response to neoadjuvant chemotherapy are included. (30) This trial was preceded by a pilot study confirming the feasibility and safety of MI-CRS in this highly selected patient subset (32) However, the investigators’ comparison of survival outcomes in this favourable-prognosis cohort with those of the broader population undergoing interval CRS may not be an ideal or meaningful comparison. The complete results of this study are awaited.

Paul Sugarbaker aptly stated ‘It is what the surgeon does not see that kills the patient’. (33) This holds particularly true for laparoscopy as it may underestimate disease volume or distribution which can be detrimental, especially in high grade malignancies like ovarian cancer and colorectal cancer. (34) Through this study we aim to quantify ‘what we cannot see’, that is the missed residual disease after minimally invasive CRS. To our knowledge this is first study of its kind which will systematically validate the oncological adequacy of MI-CRS. Only one prior study has assessed the completeness of MI-CRS, wherein all patients underwent a CT scan one month postoperatively. (35) However, the reliability of imaging in distinguishing between postoperative changes and residual disease remains limited, highlighting the need for more definitive validation.

In this study, we will employ a mini-laparotomy to thoroughly explore the abdomen and confirm the findings and adequacy of minimally invasive CRS (MI-CRS). This approach differs from most studies that rely on the completeness of cytoreduction (CC) score, which is inherently subjective. By directly verifying the surgical outcomes, this method offers a more objective and reliable validation of the quality of cytoreduction. Since a mini-laparotomy is routinely performed for specimen extraction, it is anticipated that this additional exploration will not result in any significant increase in morbidity for the patients. This is also the reason why this study has been considered an observational study. Moreover, any residual disease identified during the mini-laparotomy phase will be excised through the same incision or, if necessary, by converting to a traditional xipho-pubic laparotomy. This ensures that all patients will undergo a complete and appropriate cytoreductive surgery, with no compromise to their oncological outcomes or added risk of harm.

An additional key objective of this study is to identify the anatomical sites most frequently harbouring missed disease in MI-CRS. This information could serve as a valuable guide for operating surgeons, enabling them to better visualize and target these regions during minimally invasive procedures. Furthermore, the incidence of pathologically confirmed microscopic disease in normal-appearing target organs will be documented, underscoring the importance of their excision during cytoreductive surgery. Similarly, this study will also unravel the sites of disease which frequently led to conversion to open surgery and this knowledge may help to develop better minimally invasive surgical strategies to resect the disease. Finally, perioperative and survival outcomes of patients who undergo conversion to open surgery will be systematically recorded.

## Current status

The study is still in an early phase-11 patients have been recruited so far since 1^st^ December 2024.

## Conclusion

The MIS-CYTO study will provide crucial evidence regarding the oncological adequacy of minimally invasive CRS (MI-CRS) in the management of limited-volume peritoneal surface malignancies. Should the oncological adequacy of MI-CRS be demonstrated through this study, it will establish a strong foundation for conducting randomized trials to compare this approach with conventional CRS by laparotomy. Conversely, if it fails to demonstrate oncological adequacy, it will offer compelling justification for limiting the widespread use of MI-CRS.

## Ethics approval and consent to participate

The study was approved by the ethics committee of Shalby Cancer and Research Institute on 7^th^ October 2024 (EC/069/02)

## Consent to participate and consent publication

Written informed consent will be taken from each patient for participation in the study.

## Availability of data and materials

Not applicable

## Authors’ contributions

**Mufaddal Kazi:** conception, design, acquisition of data, analysis, interpretation, manuscript writing-original draft, manuscript-review and editing

**Vivek Sukumar:** design, interpretation, manuscript-review and editing

**Sanket Mehta:** design, interpretation, manuscript-review and editing

**Ajinkya Pawar:** design, interpretation, manuscript writing-original draft, manuscript-review and editing

**Swapnil Patel:** design, interpretation, manuscript-review and editing

**Deepti Mishra**: design, interpretation, manuscript-review and editing

**Niharika Garach:** design, interpretation, manuscript-review and editing

**Praveen Kammar:** design, interpretation, manuscript-review and editing

**Zeeba Usofi:** design, interpretation, manuscript-review and editing

**Snita Sinukumar:** design, interpretation, manuscript-review and editing

**Rajagopalan R:** design, interpretation, manuscript-review and editing

**Vivekanand Sharma:** design, interpretation, manuscript-review and editing

**Ambarish Chatterjee:** design, interpretation, manuscript-review and editing

**Rohit Ranade:** design, interpretation, manuscript-review and editing

**Aruna Prabhu:** design, interpretation, analysis, manuscript-review and editing

**Dileep Damodaran:** design, interpretation, manuscript-review and editing

**Ramakrishnan AS:** Interpretation, manuscript-review and editing

**Shigeki Kusamura:** Interpretation, manuscript-review and editing

**Avanish Saklani:** design, interpretation, manuscript-review and editing

**Kurt Van Der Speeten:** Interpretation, manuscript-review and editing

**Aditi Bhatt:** conception, design, acquisition of data, analysis, interpretation, manuscript-review and editing

## Supporting information

Reporting checklist

Study protocol

## Data Availability

All data produced in the present work are contained in the manuscript

## Acknowledgements

None

## Abbreviations used in the manuscript

CRS: cytoreductive surgery
MIS: Minimally-invasive surgery
MI-CRS: Minimally-invasive cytoreductive surgery
SL: Staging laparoscopy
HIPEC: hyperthermic intraperitoneal chemotherapy
PCI: Peritoneal cancer index
PFS: progression-free survival
RFS: relapse-free survival
OS: overall survival
CC: score-completeness of cytoreduction score

