## Supplement 1 for "MIS-CYTO: A prospective, multi-center, observational study for validation of oncological adequacy of minimally invasive cytoreductive surgery for peritoneal malignancies with limited peritoneal spread at systematic mini-laparotomy by INDEPSO"

**MIS-CYTO**

**Data collection form**

**Section 1: Demography, diagnosis, neoadjuvant treatment**

| **Demographic data and previous therapies** |
| --- |
| Name/identifier |
| Centre |
| Country |
| Age |
| Sex |
| Primary tumour site |
| For colorectal cancer-exact site of the primary |
| For ovarian cancer- exact site (ovaries, FT, peritoneum) |
| Timing of PM (synchronous/metachronous) |
| Date of diagnosis of the primary |
| Date of diagnosis of PM |
| TNM stage at diagnosis |
| FIGO stage at diagnosis (for ovarian cancer) |
| Prior CRS |
| Date of prior CRS |
| Prior CRS+HIPEC |
| Date of prior CRS+HIPEC |
| Prior surgery (not CRS) |
| Prior surgical score |
| Previous chemotherapy |
| Number of lines of previous chemotherapy |
| **Histological and molecular features** |
| Histological diagnosis |
| Tumor grade (high/low) |
| Tumor differentiation (well, moderate, poor, undifferentiated) |
| Immunohistochemistry, if relevant (positive markers) |
| Immunohistochemistry, if relevant (negative markers) |
| BRCA (mutated/wt/not evaluated) |
| KRAS (mutated/wt/not evaluated) |
| BRAF (mutated/wt/not evaluated) |
| NRAS (mutated/wt/not evaluated) |
| BAP-1 (mutated/wt/not evaluated) |
| EGFR (mutated/wt/not evaluated) |
| Ki-67 (mention exact value in percentage) |
| Any other marker (name) |
| Mutation status |
| Any other marker (name) |
| Mutation status |

| **Neoadjuvant therapies** |
| --- |
| Neoadjuvant chemotherapy |
| Intravenous/oral |
| Regimen |
| Number of cycles |
| Date of starting |
| Date of completion |
| Neoadjuvant PIPAC |
| Regimen |
| Number of applications |
| Date of start |
| Date of completion |
| Neoadjuvant port directed IP chemotherapy |
| Regimen |
| Number of cycles |
| Date of start |
| Date of completion |
| Neoadjuvant HIPEC |
| Regimen |
| Number of cycles |

**Section 2 Imaging and staging laparoscopy**

| Peritoneal MRI |
| --- |
| CT scan with oral and IV contrast |
| CT scan with IV contrast |
| PET CT scan |
| Any other (please specify) |
| **Radiological PCI** |
| **Evaluation of response** |
| RECIST criteria |
| Any other(please specify) |
| Progressive disease |
| Stable disease |
| Partial disease |
| Complete response |
| **Nodal disease** |
| Site (S) of nodal disease |
| Max diameter |
| **Ascites** |
| Ascites |
| Quantity |
| Characteristic |
| **Primary tumour site** |
| Primary tumour site (evaluated/not evaluated) |
| Disease- present or absent |
| Response in the primary |
| Regional nodes |
| Radiological T stage |
| Radiological N stage |
| Radiological M stage |
| Previous anastomosis (evaluated/not evaluated) |
| Disease- present/absent |

**Details of the radiological PCI**

| Region | Structure | Morphology (select any one) | Lesion score |
| --- | --- | --- | --- |
| 0 | Midline incision [_]  Anterior parietal peritoneum [_]  Greater omentum [_]  Transverse colon [_]  Gastrocolic ligament [_]  Transverse mesocolon [_]  Transverse colon [_] | Normal peritoneum [_]  Tumour nodule [_]  Scalloping [_]  Calcification [_]  Thickening [_]  Confluent disease [_]  Infiltration of fat tissue [_]  Omental cake [_]  Retraction [_] | 0 [_]  1 [_]  2 [_]  3 [_] |
| 1 | Right hepatic lobe surface [_]  Diaphragm [_]  Foramen of Winslow [_]  Gallbladder [_]  Hepatorenal recess [_] | Normal peritoneum [_]  Tumour nodule [_]  Scalloping [_]  Calcification [_]  Thickening [_]  Confluent disease [_]  Infiltration of fat tissue [_]  Retraction [_] | 0 [_]  1 [_]  2 [_]  3 [_] |
| 2 | Left hepatic lobe surface [_]  Lesser omentum [_]  Hepatic hilum [_]  Falciform ligament [_] | Normal peritoneum [_]  Tumour nodule [_]  Scalloping [_]  Calcification [_]  Thickening [_]  Confluent disease [_]  Infiltration of fat tissue [_]  Retraction [_] | 0 [_]  1 [_]  2 [_]  3 [_] |
| 3 | Surface of spleen [_]  Left diaphragmatic peritoneum [_]  Tail of pancreas and hilum of the spleen [_]  Anterior and posterior stomach surfaces [_] | Normal peritoneum [_]  Tumour nodule [_]  Scalloping [_]  Calcification [_]  Thickening [_]  Confluent disease [_]  Infiltration of fat tissue [_]  Retraction [_] | 0 [_]  1 [_]  2 [_]  3 [_] |
| 4 | Left colon [_]  Left paracolic gutter [_]  Left mesocolon [_] | Normal peritoneum [_]  Tumour nodule [_]  Scalloping [_]  Calcification [_]  Thickening [_]  Confluent disease [_]  Infiltration of fat tissue [_]  Retraction [_] | 0 [_]  1 [_]  2 [_]  3 [_] |
| 5 | Left pelvic peritoneum [_]  Secondary root of the mesosigmoid [_]  Sigmoid colon [_] | Normal peritoneum [_]  Tumour nodule [_]  Scalloping [_]  Calcification [_]  Thickening [_]  Confluent disease [_]  Infiltration of fat tissue [_]  Retraction [_] | 0 [_]  1 [_]  2 [_]  3 [_] |
| 6 | Uterus [_]  Left Fallopian tube [_]  Right Fallopian tube [_]  Left ovary [_]  Right ovary [_]  Bladder [_]  Pouch of Douglas [_]  Rectosigmoid junction [_] | Normal peritoneum [_]  Tumour nodule [_]  Scalloping [_]  Calcification [_]  Thickening [_]  Confluent disease [_]  Infiltration of fat tissue [_]  Retraction [_] | 0 [_]  1 [_]  2 [_]  3 [_] |
| 7 | Right pelvic peritoneum [_]  Cecum [_]  Appendix [_]  Mesoappendix [_] | Normal peritoneum [_]  Tumour nodule [_]  Scalloping [_]  Calcification [_]  Thickening [_]  Confluent disease [_]  Infiltration of fat tissue [_]  Retraction [_] | 0 [_]  1 [_]  2 [_]  3 [_] |
| 8 | Right colon [_]  Right paracolic gutter [_]  Right mesocolon [_] | Normal peritoneum [_]  Tumour nodule [_]  Scalloping [_]  Calcification [_]  Thickening [_]  Confluent disease [_]  Infiltration of fat tissue [_]  Retraction [_] | 0 [_]  1 [_]  2 [_]  3 [_] |
| 9 | Proximal jejunum [_]  Proximal mesojejunum [_] | Normal peritoneum [_]  Tumour nodule [_]  Scalloping [_]  Calcification [_]  Thickening [_]  Confluent disease [_]  Infiltration of fat tissue [_]  Mesenteric retraction [_] | 0 [_]  1 [_]  2 [_]  3 [_] |
| 10 | Distal jejunum [_]  Distal mesojejunum [_] | Normal peritoneum [_]  Tumour nodule [_]  Scalloping [_]  Calcification [_]  Thickening [_]  Confluent disease [_]  Infiltration of fat tissue [_]  Mesenteric retraction [_] | 0 [_]  1 [_]  2 [_]  3 [_] |
| 11 | Proximal ileum [_]  Proximal mesoileum [_] | Normal peritoneum [_]  Tumour nodule [_]  Scalloping [_]  Calcification [_]  Thickening [_]  Confluent disease [_]  Infiltration of fat tissue [_]  Mesenteric retraction [_] | 0 [_]  1 [_]  2 [_]  3 [_] |
| 12 | Distal ileum [_]  Distal ileum [_] | Normal peritoneum [_]  Tumour nodule [_]  Scalloping [_]  Calcification [_]  Thickening [_]  Confluent disease [_]  Infiltration of fat tissue [_]  Mesenteric retraction [_] | 0 [_]  1 [_]  2 [_]  3 [_] |
|  | Total | |  |

**Staging laparoscopy**

| Timing |
| --- |
| Before MI-CRS as a separate procedure |
| Just before commencing MI-CRS |
| Port placement |
| Sites with size |
| Number |
| Any sign of unresectability? |
| Please specify which one |

**Laparoscopic PCI**

| Region | Structure | Assessment [tick the areas assessed] | Morphology (select any one) | Lesion score |
| --- | --- | --- | --- | --- |
| 0 | Midline incision [_]  Anterior parietal peritoneum [_]  Greater omentum [_]  Omental cake [_]  Transverse colon [_]  Gastrocolic ligament [_]  Transverse mesocolon [_] | [_}  [_}  [_}  [_}  [_}  [_}  [_} | Tumour nodule [_]  Confluent nodules [_]  Plaque [_]  Thickening [_]  Scarring [_]  Adhesion [_]  Normal peritoneum [_] | 0 [_]  1 [_]  2 [_]  3 [_] |
| 1 | Right hepatic lobe surface [_]  Diaphragm [_]  Foramen of Winslow [_]  Gallbladder [_]  Hepatorenal recess [_] | [_}  [_}  [_}  [_}  [_} | Tumour nodule [_]  Confluent nodules [_]  Plaque [_]  Thickening [_]  Scarring [_]  Adhesion [_]  Normal peritoneum [_] | 0 [_]  1 [_]  2 [_]  3 [_] |
| 2 | Left hepatic lobe surface [_]  Lesser omentum [_]  Hepatic hilum/hepatoduodenal ligament [_]  Falciform ligament [_]  Superior recess of lesser sac [_] | [_}  [_}  [_}  [_} | Tumour nodule [_]  Confluent nodules [_]  Plaque [_]  Thickening [_]  Scarring [_]  Adhesion [_]  Normal peritoneum [_] | 0 [_]  1 [_]  2 [_]  3 [_] |
| 3 | Surface of spleen [_]  Left diaphragmatic peritoneum [_]  Tail of pancreas and hilum of the spleen [_]  Anterior and posterior stomach surfaces [_] | [_}  [_}  [_}  [_} | Tumour nodule [_]  Confluent nodules [_]  Plaque [_]  Thickening [_]  Scarring [_]  Adhesion [_]  Normal peritoneum [_] | 0 [_]  1 [_]  2 [_]  3 [_] |
| 4 | Left colon [_]  Left paracolic gutter [_]  Left mesocolon [_] | [_}  [_}  [_} | Tumour nodule [_]  Confluent nodules [_]  Plaque [_]  Thickening [_]  Scarring [_]  Adhesion [_]  Normal peritoneum [_] | 0 [_]  1 [_]  2 [_]  3 [_] |
| 5 | Left pelvic peritoneum [_]  Secondary root of the mesosigmoid [_]  Sigmoid colon [_] | [_}  [_}  [_} | Tumour nodule [_]  Confluent nodules [_]  Plaque [_]  Thickening [_]  Scarring [_]  Adhesion [_]  Normal peritoneum [_] | 0 [_]  1 [_]  2 [_]  3 [_] |
| 6 | Uterus [_]  Left Fallopian tube [_]  Right Fallopian tube [_]  Left ovary [_]  Right ovary [_]  Bladder [_]  Pouch of Douglas [_]  Rectosigmoid junction [_] | [_}  [_}  [_}  [_}  [_}  [_}  [_}  [_] | Tumour nodule [_]  Confluent nodules [_]  Plaque [_]  Thickening [_]  Scarring [_]  Adhesion [_]  Normal peritoneum [_] | 0 [_]  1 [_]  2 [_]  3 [_] |
| 7 | Right pelvic peritoneum [_]  Cecum [_]  Appendix [_]  Mesoappendix [_] | [_}  [_}  [_}  [_} | Tumour nodule [_]  Confluent nodules [_]  Plaque [_]  Thickening [_]  Scarring [_]  Adhesion [_]  Normal peritoneum [_] | 0 [_]  1 [_]  2 [_]  3 [_] |
| 8 | Right colon [_]  Right paracolic gutter [_]  Right mesocolon [_] | [_}  [_}  [_} | Tumour nodule [_]  Confluent nodules [_]  Plaque [_]  Thickening [_]  Scarring [_]  Adhesion [_]  Normal peritoneum [_] | 0 [_]  1 [_]  2 [_]  3 [_] |
| 9 | Proximal jejunum [_]  Proximal mesojejunum [_] | [_}  [_} | Tumour nodule [_]  Confluent nodules [_]  Plaque [_]  Thickening [_]  Scarring [_]  Adhesion [_]  Normal peritoneum [_] | 0 [_]  1 [_]  2 [_]  3 [_] |
| 10 | Distal jejunum [_]  Distal mesojejunum [_] | [_}  [_} | Tumour nodule [_]  Confluent nodules [_]  Plaque [_]  Thickening [_]  Scarring [_]  Adhesion [_]  Normal peritoneum [_] | 0 [_]  1 [_]  2 [_]  3 [_] |
| 11 | Proximal ileum [_]  Proximal mesoileum [_] | [_}  [_} | Tumour nodule [_]  Confluent nodules [_]  Plaque [_]  Thickening [_]  Scarring [_]  Adhesion [_]  Normal peritoneum [_] | 0 [_]  1 [_]  2 [_]  3 [_] |
| 12 | Distal ileum [_]  Distal ileum [_] | [_}  [_} | Tumour nodule [_]  Confluent nodules [_]  Plaque [_]  Thickening [_]  Scarring [_]  Adhesion [_]  Normal peritoneum [_] | 0 [_]  1 [_]  2 [_]  3 [_] |

**Section 3 Surgical and perioperative details**

| ASA |
| --- |
| ECOG Status |
| Date of CRS |
| Resection of the primary (Yes/No) |
| **MIS** |
| Approach- robotic or laparoscopic |
| No of PORTs used |
| Sites of PORTS with size |
| **HIPEC** |
| Open/closed |
| Drug regimen |
| Duration |
| **Surgical PCI** |
| **Perioperative outcomes** |
| Duration of surgery (MIS phase) |
| Total duration of surgery |
| Blood loss |
| **Conversion to laparotomy before CC-0/1 achieved** |
| Reason for conversion |
| Technical difficulty |
| Main area of technical difficulty |
| Time consuming |
| Excessive blood loss |
| Any other (please specify) |
| Type of anaesthesia (general or regional) |
| Epidural anaesthesia/analgesia |
| Transverse abdominis block |
| Any other (please specify) |
| Post op ventilation (hours) |
| ICU stay |
| Hospital stay |
| Grade 1-2 complications |
| 90-day grade 3-4 morbidity |
| Opioids for pain management (yes/no) |
| No of days of opioid use |
| Drains (sites and no) |
| ICD (right/left/both) |
| Day of starting oral liquid |
| Day of starting solid food |
| Time to first flatus |
| **Complications** |
| Haemorrhage |
| Specify |
| Hematological toxicity |
| Specify |
| Respiratory complications |
| Specify |
| Sepsis |
| Specify |
| Neutropenia |
| Lowest count |
| Cardiac complications |
| Specify |
| GI complications |
| Specify |
| Bowel fistula |
| Specify |
| Bowel perforation |
| Specify site |
| Renal complication |
| Specify |
| Nephrological complications |
| Specify |
| Wound dehiscence |
| Site |
| Surgical site infection |
| Return to ICU |
| Duration of stay |
| Return to operating room |
| Procedure performed/indication |
| Radiological intervention |
| Specify |
| Mortality |
| Date of death |
| Cause of death |

**Details of the surgical PCI and resections performed during MIS phase**

| Region | Structure | Morphology (select any one) | Lesion score |
| --- | --- | --- | --- |
| 0 | Midline incision [_]  Anterior parietal peritoneum [_]  Greater omentum [_]  Omental cake [_]  Transverse colon [_]  Gastrocolic ligament [_]  Transverse mesocolon [_] | Tumour nodule [_]  Confluent nodules [_]  Plaque [_]  Thickening [_]  Scarring [_]  Adhesion [_]  Normal peritoneum [_] | 0 [_]  1 [_]  2 [_]  3 [_] |
| 1 | Right hepatic lobe surface [_]  Diaphragm [_]  Foramen of Winslow [_]  Gallbladder [_]  Hepatorenal recess [_] | Tumour nodule [_]  Confluent nodules [_]  Plaque [_]  Thickening [_]  Scarring [_]  Adhesion [_]  Normal peritoneum [_] | 0 [_]  1 [_]  2 [_]  3 [_] |
| 2 | Left hepatic lobe surface [_]  Lesser omentum [_]  Hepatic hilum [_]  Falciform ligament [_] | Tumour nodule [_]  Confluent nodules [_]  Plaque [_]  Thickening [_]  Scarring [_]  Adhesion [_]  Normal peritoneum [_] | 0 [_]  1 [_]  2 [_]  3 [_] |
| 3 | Surface of spleen [_]  Left diaphragmatic peritoneum [_]  Tail of pancreas and hilum of the spleen [_]  Anterior and posterior stomach surfaces [_] | Tumour nodule [_]  Confluent nodules [_]  Plaque [_]  Thickening [_]  Scarring [_]  Adhesion [_]  Normal peritoneum [_] | 0 [_]  1 [_]  2 [_]  3 [_] |
| 4 | Left colon [_]  Left paracolic gutter [_]  Left mesocolon [_] | Tumour nodule [_]  Confluent nodules [_]  Plaque [_]  Thickening [_]  Scarring [_]  Adhesion [_]  Normal peritoneum [_] | 0 [_]  1 [_]  2 [_]  3 [_] |
| 5 | Left pelvic peritoneum [_]  Secondary root of the mesosigmoid [_]  Sigmoid colon [_] | Tumour nodule [_]  Confluent nodules [_]  Plaque [_]  Thickening [_]  Scarring [_]  Adhesion [_]  Normal peritoneum [_] | 0 [_]  1 [_]  2 [_]  3 [_] |
| 6 | Uterus [_]  Left Fallopian tube [_]  Right Fallopian tube [_]  Left ovary [_]  Right ovary [_]  Bladder [_]  Pouch of Douglas [_]  Rectosigmoid junction [_] | Tumour nodule [_]  Confluent nodules [_]  Plaque [_]  Thickening [_]  Scarring [_]  Adhesion [_]  Normal peritoneum [_] | 0 [_]  1 [_]  2 [_]  3 [_] |
| 7 | Right pelvic peritoneum [_]  Cecum [_]  Appendix [_]  Mesoappendix [_] | Tumour nodule [_]  Confluent nodules [_]  Plaque [_]  Thickening [_]  Scarring [_]  Adhesion [_]  Normal peritoneum [_] | 0 [_]  1 [_]  2 [_]  3 [_] |
| 8 | Right colon [_]  Right paracolic gutter [_]  Right mesocolon [_] | Tumour nodule [_]  Confluent nodules [_]  Plaque [_]  Thickening [_]  Scarring [_]  Adhesion [_]  Normal peritoneum [_] | 0 [_]  1 [_]  2 [_]  3 [_] |
| 9 | Proximal jejunum [_]  Proximal mesojejunum [_] | Tumour nodule [_]  Confluent nodules [_]  Plaque [_]  Thickening [_]  Scarring [_]  Adhesion [_]  Normal peritoneum [_] | 0 [_]  1 [_]  2 [_]  3 [_] |
| 10 | Distal jejunum [_]  Distal mesojejunum [_] | Tumour nodule [_]  Confluent nodules [_]  Plaque [_]  Thickening [_]  Scarring [_]  Adhesion [_]  Normal peritoneum [_] | 0 [_]  1 [_]  2 [_]  3 [_] |
| 11 | Proximal ileum [_]  Proximal mesoileum [_] | Tumour nodule [_]  Confluent nodules [_]  Plaque [_]  Thickening [_]  Scarring [_]  Adhesion [_]  Normal peritoneum [_] | 0 [_]  1 [_]  2 [_]  3 [_] |
| 12 | Distal ileum [_]  Distal ileum [_] | Tumour nodule [_]  Confluent nodules [_]  Plaque [_]  Thickening [_]  Scarring [_]  Adhesion [_]  Normal peritoneum [_] | 0 [_]  1 [_]  2 [_]  3 [_] |
|  | Total | |  |
| Details of surgical procedures performed | | | |
|  | Peritonectomy procedures | |  |
|  | Pelvic peritonectomy procedure (only region 6) | |  |
|  | Pelvic peritonectomy procedure (regions 5,6,7) | |  |
|  | Right paracolic peritonectomy procedure | |  |
|  | Left paracolic peritonectomy procedure | |  |
|  | Right anterior parietal peritonectomy procedure | |  |
|  | Left anterior parietal peritonectomy procedure | |  |
|  | Right upper quadrant peritonectomy procedure with Morrison’s pouch peritonectomy | |  |
|  | Right upper quadrant peritonectomy procedure without Morrison’s pouch peritonectomy | |  |
|  | Resection of the right diaphragm | |  |
|  | Left upper quadrant peritonectomy procedure | |  |
|  | Resection of the left diaphragm | |  |
|  | Lesser omentectomy | |  |
|  | Peritonectomy of the hepatoduodenal ligament | |  |
|  | Resection of falciform ligament | |  |
|  | Resection of umbilical round ligament | |  |
|  | Infracolic omentectomy | |  |
|  | Supracolic omentectomy with resection of the gastroepiploic arch | |  |
|  | Supracolic omentectomy with preservation of the gastroepiploic arch | |  |
|  | Glisson’s capsulectomy | |  |
|  | Small bowel mesenteric peritonectomy (1/3; >1/3: total) | |  |
|  | Peritonectomy of the superior recess of the lesser sac | |  |
|  | Peritonectomy of the foramen of Winslow | |  |
|  | Visceral resections | |  |
|  | Low anterior resection | |  |
|  | Anterior resection | |  |
|  | Sigmoidectomy | |  |
|  | Right hemicolectomy | |  |
|  | Left hemicolectomy | |  |
|  | Transverse colectomy | |  |
|  | Subtotal colectomy | |  |
|  | Total colectomy | |  |
|  | Appendicectomy | |  |
|  | Caecectomy | |  |
|  | Partial caecectomy | |  |
|  | Total hysterectomy | |  |
|  | Right and/or left salpino-oophorectomy | |  |
|  | Splenectomy | |  |
|  | Cholecystectomy | |  |
|  | Wedge resection of liver | |  |
|  | Any other liver resection (specify) | |  |
|  | Distal gastrectomy | |  |
|  | Subtotal gastrectomy | |  |
|  | Total gastrectomy | |  |
|  | Distal pancreatectomy | |  |
|  | Small bowel resection | |  |
|  | Site of resection | |  |
|  | Length of resected segment | |  |
|  | Any other visceral resection (please specify) | |  |
|  | Regional lymphadenectomy | |  |
|  | Pelvic (right/left/ bilateral) | |  |
|  | Para-aortic (mention upper limit) | |  |
|  | D2 lymphadenectomy with gastrectomy | |  |
|  | Mesocolic lymphadenectomy (with colonic resection) | |  |
|  | Any other (please specify) | |  |
|  | No of bowel anastomoses | |  |
|  | Sites of bowel anastomosis | |  |
|  | Type of anastomosis  Hand sewn  Stapled | |  |
|  | Diverting stomy | |  |
|  | Site | |  |
|  | Temporary/ permanent | |  |
|  | Delivery of MIS specimen | |  |
|  | Through the vagina | |  |
|  | Through the laparotomy incision | |  |

**Section 4 Systematic laparotomy phase**

**Residual surgical PCI after MI-CRS and details of systematic laparotomy**

| **Region** | **Exact site of residual disease** | **Morphology (select any one)** | **Lesion score** |
| --- | --- | --- | --- |
| 0 |  |  | 0 [_] 1 [_]  2 [_] 3 [_] |
| 1 |  |  | 0 [_] 1 [_]  2 [_] 3 [_] |
| 2 |  |  | 0 [_] 1 [_]  2 [_]3 [_] |
| 3 |  |  | 0 [_]1 [_]  2 [_] 3 [_] |
| 4 |  |  | 0 [_] 1 [_]  2 [_] 3 [_] |
| 5 |  |  | 0 [_] 1 [_]  2 [_] 3 [_] |
| 6 |  |  | 0 [_] 1 [_]  2 [_] 3 [_] |
| 7 |  |  | 0 [_] 1 [_]  2 [_] 3 [_] |
| 8 |  |  | 0 [_] 1 [_]  2 [_] 3 [_] |
| 9 |  |  | 0 [_] 1 [_]  2 [_] 3 [_] |
| 10 |  |  | 0 [_] 1 [_]  2 [_] 3 [_] |
| 11 |  |  | 0 [_] 1 [_]  2 [_] 3 [_] |
| 12 |  |  | 0 [_] 1 [_]  2 [_] 3 [_] |
|  | Total | |  |
| **Details of surgical procedures performed** | | | |
|  | **Peritonectomy procedures** | |  |
|  | Pelvic peritonectomy procedure (only region 6) | |  |
|  | Pelvic peritonectomy procedure (regions 5,6,7) | |  |
|  | Right paracolic peritonectomy procedure | |  |
|  | Left paracolic peritonectomy procedure | |  |
|  | Right anterior parietal peritonectomy procedure | |  |
|  | Left anterior parietal peritonectomy procedure | |  |
|  | Right upper quadrant peritonectomy procedure with Morrison’s pouch peritonectomy | |  |
|  | Right upper quadrant peritonectomy procedure without Morrison’s pouch peritonectomy | |  |
|  | Resection of the right diaphragm | |  |
|  | Left upper quadrant peritonectomy procedure | |  |
|  | Resection of the left diaphragm | |  |
|  | Lesser omentectomy | |  |
|  | Peritonectomy of the hepatoduodenal ligament | |  |
|  | Resection of falciform ligament | |  |
|  | Resection of umbilical round ligament | |  |
|  | Infracolic omentectomy | |  |
|  | Supracolic omentectomy with resection of the gastroepiploic arch | |  |
|  | Supracolic omentectomy with preservation of the gastroepiploic arch | |  |
|  | Glisson’s capsulectomy | |  |
|  | Small bowel mesenteric peritonectomy (1/3; >1/3: total) | |  |
|  | Peritonectomy of the superior recess of the lesser sac | |  |
|  | Peritonectomy of the foramen of Winslow | |  |
|  | Resection of previous surgical scar | |  |
|  | Resection of PORT sites | |  |
|  | **Visceral resections** | |  |
|  | Low anterior resection | |  |
|  | Anterior resection | |  |
|  | Sigmoidectomy | |  |
|  | Right hemicolectomy | |  |
|  | Left hemicolectomy | |  |
|  | Transverse colectomy | |  |
|  | Subtotal colectomy | |  |
|  | Total colectomy | |  |
|  | Appendicectomy | |  |
|  | Caecectomy | |  |
|  | Partial caecectomy | |  |
|  | Total hysterectomy | |  |
|  | Right and/or left salpingo-oophorectomy | |  |
|  | Splenectomy | |  |
|  | Cholecystectomy | |  |
|  | Wedge resection of liver | |  |
|  | Any other liver resection (specify) | |  |
|  | Distal gastrectomy | |  |
|  | Subtotal gastrectomy | |  |
|  | Total gastrectomy | |  |
|  | Distal pancreatectomy | |  |
|  | Small bowel resection | |  |
|  | Site of resection | |  |
|  | Length of resected segment | |  |
|  | Any other visceral resection (please specify) | |  |
|  | **Regional lymphadenectomy** | |  |
|  | Pelvic (right/left/ bilateral) | |  |
|  | Para-aortic (mention upper limit) | |  |
|  | D2 lymphadenectomy with gastrectomy | |  |
|  | Mesocolic lymphadenectomy (with colonic resection) | |  |
|  | Any other (please specify) | |  |
|  | **No of bowel anastomoses** | |  |
|  | Sites of bowel anastomosis | |  |
|  | Type of anastomosis  Hand sewn  Stapled | |  |
|  | **Diverting o’stomy** | |  |
|  | Site | |  |
|  | Temporary/ permanent | |  |

**Section 5 Histopathological assessment**

| **Frozen section** |
| --- |
| Regions sent |
| Disease (yes/no) |
| **MI-CRS phase** |
| List peritoneal regions submitted |
| List regions positive for malignancy |
| List viscera submitted |
| List viscera positive for malignancy |
| List regional lymph nodes dissected |
| List sites of positive regional nodes |
| Presence of disease in target regions |
| Greater omentum |
| Lesser omentum |
| Falciform ligament |
| Umbilical round ligament |
| Port sites |
| **Laparotomy phase** |
| List peritoneal regions submitted |
| List regions positive for malignancy |
| List viscera submitted |
| List viscera positive for malignancy |
| List regional lymph nodes submitted |
| List sites of positive regional nodes |
| Presence of disease in target regions |
| Greater omentum |
| Lesser omentum |
| Falciform ligament |
| Umbilical round ligament |
| Abdominal scar |
| **Pathological response to neoadjuvant chemotherapy** |
| Bohm score(for ovarian cancer) |
| Response grade |
| PRGS |
| Response grade |
| Any other score used |
| Grade |
| Additional comments |
| Pathological complete response (y/n) |
| **Evaluation of the primary tumour** |
| Disease in the primary |
| Maximum tumour diameter |
| T-stage |
| N-stage |
| No of dissected nodes |
| No of positive nodes |
| Extracapsular spread |
| Lymphovascular invasion |
| Perineural invasion |
| Immunohistochemistry-positive markers |
| Immunohistochemistry-negative markers |

**Section 6 Adjuvant therapy and follow up**

| Adjuvant therapy |
| --- |
| Adjuvant therapy planned |
| Date of start of adjuvant therapy |
| Chemotherapy regimen |
| Number of cycles |
| Targetted therapy |
| Drug |
| Number of cycles |
| Date of last cycle |
| Maintenance therapy |
| Drug |
| Duration |
| Follow-up |
| Last follow-up |
| Disease status |
| Date of diagnosis of recurrence |
| Site of recurrence |
| Site of metastases |
| Treatment for recurrence/metastatic disease |
| Date of death |
| Cause of death |
