## Supplementary material for "MIS-CYTO: A prospective, multi-center, observational study for validation of oncological adequacy of minimally invasive cytoreductive surgery for peritoneal malignancies with limited peritoneal spread at systematic mini-laparotomy by INDEPSO": Study protocol

**Clinical Investigation Plan**

Trial registration: **CTRI/2024/11/076312** (Clinical Trials Registry of India; subsidiary of clinicaltrials.gov)

| **SPONSOR** | **CRO REPRESENTATIVE** |
| --- | --- |
| **SOCIETY OF PERITONEAL SURFACE ONCOLOGY (SPSO), India**  4th Floor Dalawal chambers, JB road Nariman Point, Mumbai 400021 | **None** |

**Confidentiality Notice**

This document contains confidential information about **Society of Peritoneal Surface Oncology, India**. This document must not be disclosed to anyone other than study staff and members of the Independent Ethics Committee. The information in this document cannot be used for any purpose other than the conduct or evaluation of the clinical trial without the prior written consent of **Society of Peritoneal Surface Oncology, India** which is the registered society for INDEPSO **(Indian Network for Development of Peritoneal Surface Oncology)**

**Clinical Investigation Plan - Cover Page**

| **Title of Study:** | MIS-CYTO: Prospective multi-center validation of oncological adequacy of MInimally invaSive CYTOreductive surgery for peritoneal malignancies with limited peritoneal spread at systematic mini- laparotomy |
| --- | --- |
| **Study Design:** | This study aims to validate minimally invasive cytoreductive surgery (MI-CRS )for limited peritoneal cancer spread by demonstrating the oncological adequacy of MI-CRS compared to cytoreductive surgery (CRS) performed by laparotomy  All patients will undergo systematic mini-laparotomy after MI-CRS.  This is a single-arm, prospective, interventional study. Patients with both primary and secondary peritoneal malignancies eligible for CRS will be included in the study if the peritoneal cancer index (PCI) is less than 10 and imaging shows no signs of unresectability. Patients with a low PCI (less than 10) but in whom a CC-0/1 resection is not possible will be excluded from the study. Patients with a low PCI on imaging in whom minimally-invasive surgery (MIS) is not feasible like those with prior CRS by laparotomy, prior multiple and extensive laparotomies and those in whom the MIS approach is contraindicated will not be included in the study but the data will be captured. This will provide information on how many patients with a low PCI were not eligible for MI-CRS. A staging laparoscopy will be performed for all patients just prior to the MI-CRS or as a separate intervention as permissible. The laparoscopic findings will be recorded in the prespecified format with of each region of the peritoneal cavity and submitted to the regulatory committee. If laparoscopy reveals any sign of unresectable disease, these patients will be excluded from the study but data will be collected at baseline. There will be no restriction on the sites of disease that could be resected provided the surgeon is comfortable performing the procedure by the minimally invasive (MI) approach. MI-CRS could be performed by the laparoscopic or robotic approach. At the end of the procedure, a video of the sites of resection demonstrating the completeness of resection will be submitted to the regulatory committee for each patient. The video should be shot in a prespecified format. At the end of the MI-CRS, when a CC-0/1 resection is achieved, the abdomen will be explored through a mini-laparotomy incision. This incision should extend from mid-way between the xiphoid and umbilicus to midway between the umbilicus and pubic symphysis. An-arch preserving complete greater omentectomy will be performed for all patients if not performed during MI-CRS. The falciform ligament and the umbilical round ligament will be resected for all patients. Further exploration will be performed as described in section 7.3.1. All resected tissues will be submitted for pathological evaluation. If hyperthermic intraperitoneal chemotherapy (HIPEC) is indicated, it will be performed after the open part of the procedure is completed by the open or the closed method as is the practice of the surgeon. Patients in whom the surgeon decides to convert to laparotomy to obtain a complete resection will be excluded from the study. These patients will, however, continue to be followed-up. The reason for conversion will be clearly stated. All the additional procedures (resection of the greater omentum, falciform and umbilical round ligaments) will be performed in patients who conversion to laparotomy before a CC-0/1 resection is achieved. The postoperative management and pre and postoperative oncological treatments will be as deemed appropriate by the treating physician/institution. The additional disease sites that were identified by the surgeon will be studied. Post-operative outcomes will be recorded and include the ICU-stay, hospital stay, grade 3-4 complications at 30 and 90-days, grade 1-2 complications, 90-day mortality, time-to-return of bowel function. The perioperative pain management will be as per the existing institutional protocol but will be captured in detail. |
| **Type of Study:** | Observational, prospective, non-randomized, and multi-centre study. |
| **Indication:** | All adult patients with peritoneal metastasis of ovarian, colorectal, and gastric cancer; pseudomyxoma peritonei or mesothelioma, with limited disease extent (PCI<10) who are eligible for minimally-invasive cytoreductive surgery |
| **Study Period:** | 15^th^ November 2024  14^th^ November 2027 |
| **Principal Investigator with designation and departments:** | **Dr. Aditi Bhatt**  Sr. consultant Surgical Oncologist,  Shalby Cancer and Research Institute, Ahmedabad  India-380015   **Dr. Sanket Mehta**  Director, Specialty Surgical Oncology Hospital and Research Centre, Ghatkopar, Mumbai  |
| **Co-Investigator with designation and departments:** | 1. **Dr. Mufaddal Kazi**   Associate Professor, Surgical Oncology  Department of Colorectal and gastrointestinal surgery,  Tata Memorial Hospital, Parel, Mumbai    2. **Dr. Swapnil Patel**   Surgical Oncologist  Clinical Lead - GI, HPB & Peritoneal Surface Oncology  Medical Director, Upkar Cancer Institute (unit of Upkar Hospital Pvt Ltd) Varanasi    3. **Dr. Deepti Mishra**   Consultant Surgical oncology,  Thangam cancer centre, Namakkal, Tamil Nadu    4. **Dr. Dileep Damodaran**   Consultant, Surgical Oncology,  MVR cancer centre and research institute,  Calicut, India  |
| **Sponsor*:** | SPSO |
| **Name and Address of the Investigation Site:** | Shalby Hospital, SG highway, Ahmedabad-380015; India  Specialty Surgical Oncology Hospital and Research Centre, Ghatkopar, Mumbai |
| **Standard and Regulatory reference** |  |

**Table of Contents**

1. Introduction

1. **Introduction**

Cytoreductive surgery (CRS) which is the cornerstone of potential curative or life-prolonging treatment for peritoneal malignancies (PM) is conventionally performed by laparotomy.[1,2] A midline, xiphoid-to-pubis incision is employed to explore the whole abdomen and perform peritonectomies and visceral resections as required to achieve a complete cytoreduction (CC-0 resection).[2] With an increase in awareness about PM and development of imaging techniques and protocols, peritoneal malignancy is detected early in many patients when the spread is limited to few regions of the peritoneal cavity.[2] Minimally-invasive surgery (MIS) has gained increasing acceptance for the treatment of primary gastrointestinal and gynecological malignancies with the demonstration of non-inferiority to open surgery in well-designed clinical studies and/or randomized clinical trials.[3,4,5,6,7] Surgeons have explored the possibility of minimally invasive CRS (MI-CRS) for patients with limited peritoneal cancer spread. The early results show that the procedure is feasible.[8] While some surgeons recommend it only for low grade malignancies with a low peritoneal cancer index (PCI) (PCI<10) like low grade pseudomyxoma peritonei (PMP) and multi-cystic peritoneal mesothelioma, others have recommended it for high-grade malignancies too.[9,10,11] All the available evidence comes from retrospective cohort studies.[9,10,11] One multi-institution study found an increase in recurrence in patients with high grade malignancies while another large study found no increase in early recurrence.[9,12] However, the follow-up in the latter study was short (median follow up of 12 months) and the number of patients for each primary site low to determine the oncological safety of the procedure.[12] There are two randomized trials comparing minimally invasive and open CRS, the Mirrors trial and the LANCE trial for interval CRS in advanced ovarian cancer.[13,14] In the LANCE trial only patients that have a complete or near complete response to neoadjuvant chemotherapy are included.[13] This trial was preceded by a pilot study showing the feasibility and safety of MI-CRS in this subset of patients.[15] However, the investigators compare the survival outcomes in a cohort of patients with extremely good prognosis to that achieved in the whole cohort of patients undergoing interval CRS might not be an ideal comparison.

Before such a trial is conducted for other malignancies, the technical feasibility needs to be demonstrated. There are two different kinds of technical challenges- performing peritonectomy procedures and visceral resections employing the minimally invasive approach is technically challenging and there are few formal descriptions of the techniques of MI-CRS. The second challenge is assessing the disease extent and residual disease accurately.

There are quality indicators to for oncological surgeries that include the surgeons’ assessment of completeness of surgery and pathological evaluation of surgical specimens.[16,17] For CRS, the only metric to evaluate the quality of surgery is the surgeon’s documentation of completeness of cytoreduction that is based on the intraoperative assessment of residual disease on visual inspection and palpation of intra-abdominal structures/organs. This is one of the challenges of MI-CRS as there are is no tactile feedback. Palpation of bowel, omenta and other regions which might reveal peritoneal deposits is lacking. It may not be possible to visualize some regions as clearly as in open surgery like the posterior edge of the spleen, the sub-pyloric region, and even the lesser omentum.

When comparing the MI and open approaches for other surgical procedures, the surgery that is performed in largely dependent on preoperative imaging findings. When the surgery is dependent on intraoperative exploration, then feasibility and adequacy of the MI approach needs to be demonstrated by converting the procedure to open surgery to confirm the findings of the MI approach.[18]

This study is a prospective validation study in which the adequacy and safety of the minimally invasive CRS will be confirmed by performing a min-laparotomy at the end of the procedure and systematically exploring the abdominal cavity and resecting areas of disease, if any. A mini-laparotomy is generally performed at the end of MI-CRS to deliver the specimen.

**Rationale**

The ‘gold standard’ for assessment of the presence or absence of peritoneal disease is laparotomy. The main concern in MI-CRS is missing disease in regions that were difficult to access by MIS or when the tumor nodules are embedded in fat tissue and cannot be visualized unless palpation is performed. Though imaging can detect such nodules, it has the limitation of missing nodules that measure less than 5mm.

There is no information in literature about the sites of peritoneal disease that can or cannot be resected by the minimall-invasive approach. This is particularly a concern for regions like the upper abdomen and the small bowel mesentery. The design of this study will enable us to provide crucial information on the sites that are resected during MI-CRS and the quality of the resection.

There will be no additional intervention in the study since a mini-laparotomy is performed to deliver the specimen of cytoreductive surgery. Even in cases when the vagina is opened it may be difficult to deliver the specimen through the vagina without extensive handling which poses the risk of tumor spillage.

**Novelty**

This is the first study in which we will perform a systematic exploration of the abdominal cavity via mini- laparotomy after the MI resection for all patients undergoing MI-CRS to confirm the oncological adequacy of MI-CRS. Only one previous study has evaluated the completeness of MI-CRS and a CT scan was performed in all patients 1 month after the MI-CRS.[19] There are limitations of imaging in distinguishing postoperative changes from residual disease. All other studies use the CC-score which is a subjective assessment performed by the surgeon during MI-CRS. As described above, some areas could be difficult to assess during the staging procedure. There is a lack of tactile feedback which is present in open surgery and allows detection of nodules that could be missed due to their anatomical location or being completely embedded in fat as with nodules in the mesocolon or in the appendices epiploicae.

There is a specific format for documenting the findings of the staging laparoscopy. Laparoscopic assessment could also vary among surgeons and this format could help to bring about some uniformity in exploration and documentation of the findings.

If the oncological adequacy of MI-CRS is established, then there will be a strong basis for performing randomized trials to compare this approach with CRS by laparotomy. If the oncological adequacy cannot be demonstrated, then this will provide a strong basis for restricting the widespread use of MI-CRS.

### **2. Identification of the clinical investigation plan**

| **Title of the clinical investigation:** | MIS-CYTO: Prospective multi-center validation of oncological adequacy of MInimally invaSive CYTOreductive surgery for peritoneal malignancies with limited peritoneal spread at systematic mini-laparotomy |
| --- | --- |
| **Reference number identifying the specific clinical investigation:** | **CTRI/2024/11/076312** |

#### **2.1. Abbreviations and acronyms**

| CRS | Cytoreductive Surgery |
| --- | --- |
| HIPEC | Hyperthermic (or Heated) Intraperitoneal Chemoperfusion |
| MIS | Minimally invasive surgery |
| MI-CRS | Minimally invasive cytoreductive surgery |
| CRF | Case Report Form |
| PCI | Peritoneal cancer index |
| CC-score | Completeness of cytoreduction score |
| OS | Overall survival |
| PFS | Progression-free survival |
| ECOG | Eastern Cooperative Oncology Group |
| ASA | American Society of Anesthesiologists |
| CIP | Clinical Investigation Plan |
| rPCI | Radiological Peritoneal Cancer Index |
| ITT | Intention to treat analysis |
| AE | Adverse Event |
| PP | Per-protocol Population |
| DMC | Data Monitoring Committee |
| EC | Ethics Committee |
| GCP | Good Clinical Practice |
| PMP | Pseudomyxoma Peritonei |
| PeM | Peritoneal Mesothelioma |
| ICF | Informed Consent Form |
| PI | Principal Investigator |
| SAE | Serious Adverse Event |
| SDV | Source Data Verification |

### 3. **Sponsor details**

| **SI. No.** | **Name** | **Address** | **Scope** |
| --- | --- | --- | --- |
| 1 | Society of Peritoneal Surface Oncology, India | 4th Floor Dalawal chambers, JB road Nariman Point, Mumbai 400021 | The administrative cost will be borne by SPSO |

### **4. Investigator and site details**

| **Principal Investigator:** | **1. Dr. Aditi Bhatt**  Sr consultant Surgical Oncologist,  Shalby Cancer and Research Institute,  Ahmedabad, India-380015    2. **Dr. Sanket Mehta**   Director, Specialty Surgical Oncology Hospital and Research Centre, Ghatkopar, Mumbai-400004  |
| --- | --- |
| **Co-Investigator:** | 3. **Dr. Mufaddal Kazi**   Associate Professor, Surgical Oncology  Department of Colorectal and gastrointestinal surgery,  Tata Memorial Hospital, Parel, Mumbai    4. **Dr. Swapnil Patel**   Surgical Oncologist  Clinical Lead - GI, HPB & Peritoneal Surface Oncology  Medical Director, Upkar Cancer Institute (unit of Upkar Hospital Pvt Ltd) Varanasi    5. **Dr. Deepti Mishra**   Consultant Surgical oncology,  Thangam cancer centre, Namakkal, Tamil Nadu    6. **Dr. Dileep Damodaran**   Consultant, Surgical Oncology,  MVR cancer centre and research institute,  Calicut, India  |
| **Investigation site:** | 1. Shalby Hospital, SG highway, Ahmedabad-380015; India 2. Specialty Surgical Oncology Hospital and Research Centre, Ghatkopar, Mumbai |
| **Names and addresses of external organizations:** | 1. Tata Memorial Hospital, Parel, Mumbai 2. Upkar Cancer Institute (unit of Upkar Hospital Pvt Ltd) Varanasi, India 3. Thangam cancer center, Namakkal, India 4. MVR cancer centers and research institute, Calicut, India |

### **5. Overall synopsis of the clinical investigation**

| **Number of subjects:** | 100 patients will be enrolled in the study over a period of 3 years.  The sample size calculation is discussed in detail later in this document |
| --- | --- |
| **Duration of the clinical investigation:** | The duration of the clinical investigation is 3 years months i.e., from 15^th^ November 2024 to 14^th^ November 2027. |
| **Follow-up:** | The follow-up of the subjects will be conducted for 5 years from the date of intervention. |

### **6. Objectives and hypotheses of the clinical investigation**

#### **6.1. Goal of the study**

The to validate MI-CRS for limited peritoneal cancer spread by demonstrating the oncological adequacy of MI-CRS compared to CRS performed by laparotomy.

#### **6.2. Objective**

**Primary Objective:**

To demonstrate that MI-CRS performed in well selected patients leads to a low incidence of missed peritoneal disease (<5%)

**Secondary Objectives:**

To study the rate of conversion to laparotomy to achieve a complete cytoreduction and the reasons for conversion

To study the sites where disease was missed during MI-CRS

To study which peritonectomies/resections can be performed during MI-CRS

To study the perioperative and survival outcomes after MI-CRS

#### **6.3. Hypothesis**

MI-CRS for limited peritoneal cancer spread is has the same oncological outcomes as CRS performed by laparotomy in patients with low PCI and those with high-grade disease too.

#### **6.4. Risks and anticipated adverse device effects that are to be assessed**

There are two main concerns- inability to remove the tumour completely by the MI-CRS approach. A conversion to xipho-pubic laparotomy is permissible if the surgeon cannot remove all the disease by minimally-invasive surgery.

The second concern is missing disease that was not visualized during MI-CRS. A mini-laparotomy will be performed in all patients, which should negate the chances of missed disease.

The other adverse effects are related to CRS itself on which MI-CRS should not have an impact.

Specific to MIS, injury to viscera during dissection or vascular structures could occur. The incidence of these should be less than 1%. The adverse events specific to MIS will be monitored on a yearly basis by a regulatory committee that will review the all the documents and videos of each patient enrolled to the study.

### **7. Design of the clinical investigation**

#### **7.1. General design of the clinical investigation**

This is a single-arm, prospective, observational study. Patients with both primary and secondary peritoneal malignancies eligible for CRS will be included in the study if the PCI<10 and imaging shows no signs of unresectability. Patients with a low PCI (<10) but in whom a CC-0/1 resection is not possible will be excluded from the study. Patients with a low PCI on imaging in whom MIS is not feasible like those with prior CRS by laparotomy, prior multiple and extensive laparotomies and those in whom the MIS approach is contraindicated will not be included in the study but the data will be captured. This will provide information on how many patients with a low PCI were not eligible for MI-CRS. A staging laparoscopy will be performed for all patients just prior to the MI-CRS or as a separate intervention as permissible. The laparoscopic findings will be recorded in the prespecified format with of each region of the peritoneal cavity and submitted to the regulatory committee. Each of the three members of regulatory committee will assess the quality of the video independently **(Annexure 1)**. If laparoscopy reveals any sign of unresectable disease, these patients will be excluded from the study but data will be collected at baseline. There will be no restriction on the sites of disease that could be resected provided the surgeon is comfortable performing the procedure by the MI approach. MI-CRS could be performed by the laparoscopic or robotic approach. At the end of the procedure, a video of the sites of resection demonstrating the completeness of resection will be submitted to the regulatory committee for each patient. Each of the three members of regulatory committee will assess the quality of the video independently **(Annexure 2)**. At the end of the MI-CRS, when a CC-0/1 resection is achieved, the abdomen will be explored through a midline mini-laparotomy that extends from mid-way between the xiphoid process to the umbilicus to mid-way between the umbilicus and pubic symphysis . The surgical protocol should be followed for the staging laparoscopy, MI-CRS phase and mini-laparotomy phase **(7.3.1)**. All resected tissues will be submitted for pathological evaluation. If HIPEC is indicated, it will be performed after the open part of the procedure is completed by the open or the closed method as is the practice of the surgeon. Patients in whom the surgeon decides to convert to laparotomy to obtain a complete resection will undergo laparotomy and will be excluded from the study but all the data will be collected. The reason for conversion will be clearly stated. All the above additional procedures will be performed in patients who conversion to laparotomy before a CC-0/1 resection is achieved. The study design is elaborated in **Figure 1**.

Figure 1: Study design

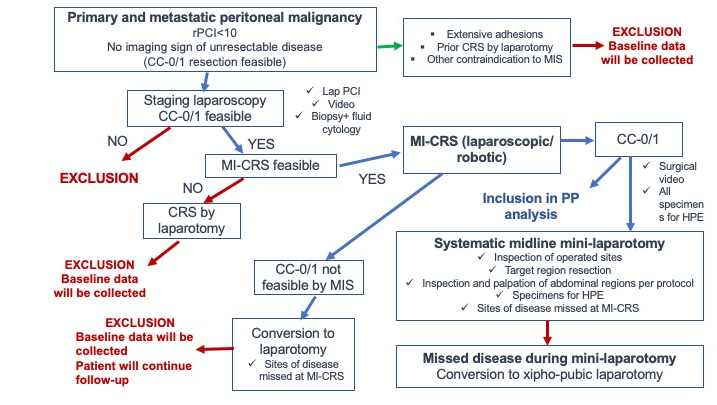

##### **7.1.1. Endpoint**

**Primary end-point**

- Proportion of patients with pathologically proven peritoneal disease that was not identified/resected during MI-CRS

**Secondary end-points**

- Incidence of pathologically proven disease in ‘normal appearing’ target regions (greater omentum, lesser omentum, falciform ligament and/or umbilical round ligament) resected during laparotomy which were not resected during MI-CRS
- Duration of hospitalization
- Grade 3-4 morbidity at 90 days
- Perioperative outcomes : operative time
- Perioperative outcomes : blood loss
- Perioperative outcomes: use of opioids
- Recovery of bowel function (time to first flatus)
- 1-year PFS
- 3-year PFS and OS
- 5-year PFS and OS
- Rate of conversion to laparotomy

##### **7.1.2.** **Methods and timing for assessing, recording, and analyzing variables**

After all the preliminary work up and investigations, patients who meet the inclusion criteria will be included. Staging laparoscopy will be performed for all patients just prior to the MI-CRS or as a separate intervention as permissible. The laparoscopic findings will be recorded in the prespecified format with of each region of the peritoneal cavity and submitted to the regulatory committee. If laparoscopy reveals any sign of unresectable disease, these patients will be excluded from the study but data will be collected at baseline. There will be no restriction on the sites of disease that could be resected provided the surgeon is comfortable performing the procedure by the MI approach. MI-CRS could be performed by the laparoscopic or robotic approach. At the end of the procedure, a video of the sites of resection demonstrating the completeness of resection will be submitted to the regulatory committee for each patient. The video should be shot in a prespecified format. At the end of the MI-CRS, when a CC-0/1 resection is achieved, the abdomen will be explored through a midline mini-laparotomy that extends from mid-way between the xiphoid process to the umbilicus to mid-way between the umbilicus and pubic symphysis. An-arch preserving complete greater omentectomy will be performed for all patients if not performed during MI-CRS. The falciform ligament and the umbilical round ligament will be resected for all patients. Further exploration will be performed as described below. All resected tissues will be submitted for pathological evaluation. If HIPEC is indicated, it will be performed after the open part of the procedure is completed by the open or the closed method as is the practice of the surgeon. Patients in whom the surgeon decides to convert to laparotomy to obtain a complete resection will undergo laparotomy and will be excluded but these patients with continue follow-up and all their data will be collected. The reason for conversion will be clearly stated by the chief surgeon. All the above additional procedures will be performed in patients whom conversion to laparotomy is performed before a CC-0/1 resection is achieved. The postoperative management and pre and postoperative oncological treatments will be as deemed appropriate by the treating physician/institution. The additional disease sites that were identified by the surgeon will be studied. Post-operative outcomes will be recorded and include the ICU-stay, hospital stay, grade 3-4 complications at 30 and 90-days, grade 1-2 complications, 90-day mortality, time-to-return of bowel function. The perioperative pain management will be as per the existing institutional protocol but will be captured in detail. The study design is elaborated in Figure 1.

##### **7.1.3. Definition of completion of the clinical study**

The completion of a clinical investigation shall be deemed to coincide with the last visit of the last subject and when follow-up is complete for the clinical investigation, whether the clinical investigation concluded according to the pre-specified clinical investigation plan or was terminated prematurely unless another point in time for such end is set out in the clinical investigation plan.

#### **7.2. Subjects**

##### **7.2.1. Inclusion Criteria**

- Age between 18-75 years
- ECOG performance status 1 or less than 1, ASA 1 or 2
- Patients with biopsy proven peritoneal malignancy that includes the following primary sites: colorectal cancer, epithelial ovarian cancer, peritoneal mesothelioma, pseudomyxoma peritonei, gastric cancer with radiological PCI<10 and no sign of unresectability on imaging
- Patients with advanced ovarian cancer undergoing primary or interval cytoreductive surgery will be included if the radiological PCI is less than 10
- Patients with recurrent ovarian cancer with 1-2 tumor sites will be included if MI-CRS is feasible
- Laparoscopic PCI<10
- Age 18-70 years
- Normal blood counts and blood biochemistry
- No medical contraindication to major abdominal surgery
- Patients who have had previous laparoscopic surgery for primary tumor resection will be included if all the other criteria are fulfilled
- Patients with prior laparotomy could be included if the same was not performed for treating peritoneal malignancy by cytoreductive surgery except selected cases of recurrent ovarian cancer with only 1-2 sites of disease.

##### **7.2.2. Exclusion Criteria**

- Prior CRS+/- HIPEC
- Patients with a PCI<10 and imaging features indicate that CC-0/1 is not possible
- Patients with PCI<10 in whom unresectable disease is discovered at staging laparoscopy
- Patients undergoing prophylactic procedures/second look procedures
- Age <18 years or >70 years
- Primary tumor site excluding colorectal cancer, gastric cancer, advanced ovarian cancer, peritoneal mesothelioma and pseudomyxoma peritonei
- Patient refusing consent
- Renal impairment, defined as glomerular filtration rate (GFR) less than 40 mL/min
- Impaired liver function defined as bilirubin ≥ 1.5 × UNL (upper normal limit) and transaminases above the upper limit of normal range
- ASA III and above (ASA grade III: A patient with a severe systemic disease that is not life-threatening)
- Patients with distant organ metastases like liver and lung metastases, extra abdominal metastasis
- Patients not fit for laparoscopy
- Patients with small or large bowel obstruction/perforation or fistula

##### **7.2.3. Criteria and procedures for subject withdrawal or lost to follow-up**

Each participant has the right to withdraw from the study at any time by with or without providing a reason. In addition, the investigator may discontinue a participant from the study at any time if the investigator considers it necessary for any reason including:

- Significant protocol deviation (MI-CRS not feasible)

- Consent withdrawn

The subject can withdraw or discontinue from the study if there are any side effects or adverse events that occurred during the study or if the study is affecting the health of the subject. The subject is free to withdraw from the study at any time after giving a reason or with no reason. The decision to continue or to withdraw will be concluded based on the investigator’s advice and the reason for withdrawal will be noted in the CRF.

##### **7.2.4. Point of enrolment**

Written informed consent has to be provided by the subject for their enrolment in the clinical trial.

Subjects must be able to understand and comply with written and verbal protocol requirements, instructions, and protocol-stated restrictions.

Subjects should be able to sign and date the written informed consent document acknowledging their desire to participate in the study, or if unable to sign, a legal representative consent has to be obtained before initiating the clinical trial.

##### **7.2.5. Total expected duration of the clinical investigation**

The total expected duration of the clinical investigation is 8 years.

##### **7.2.6. Number of subjects and their distribution of enrolment**

100 patients will be enrolled

##### **7.2.7. Enrolment period**

The time for enrollment of 100 patients is 3 years

#### **7.3. Procedures**

##### **7.3.1. Description of all the clinical investigation-related procedures**

**Surgical intervention**

The procedure is performed under general anesthesia.

The patient selection for surgery, the preoperative work up, perioperative and postoperative management and additional oncological treatments will be according to the protocols followed at each center. In the preoperative work up, the radiological PCI should be documented in the prescribed format.

**Staging laparoscopy**

- The staging laparoscopy is performed as an independent procedure or just prior to commencing with the MI-CRS.
- All the ports for this procedure should be placed in the midline when feasible.
- The 13-region PCI should be evaluated. The PCI will be reported using the form provided that will include a description of the morphology of the lesion.
- A video showing all the regions explored in the laparoscopic assessment should be shot and submitted with the clinical data to the regulatory committee.
- This should include sites that have overt disease, areas of suspicion and uninvolved regions as well.
- Peritoneal biopsies will be taken to confirm the presence of peritoneal malignancy if not done already.
- Ascitic fluid will be sampled or peritoneal washings after instillation of 200 ml of normal saline will be taken and sent for cytological and/or cell block analysis.

**MI-CRS**

- This procedure could be performed laparoscopically or robotically.
- The port placement is at the discretion of the surgeon.
- A staging laparoscopy will be performed as described above if it was not performed earlier. The procedure begins with reconfirmation of the previous laparoscopic findings.
- CRS is then performed to achieve a CC-0 resection.
- Frozen section is permitted to aid decision making
- The technique of peritonectomy and visceral resections will be according to the preference and comfort of the surgeon.
- Some surgeons prefer to perform a mini-laparotomy to assess the small bowel and its mesentery during MI-CRS but this is not permitted in the MI-CRS phase of this study. The main reason is to document the incidence of missed disease if the small bowel mesentery is not assessed by a mini-laparotomy
- The specimens should be delivered when the mini-laparotomy is performed.
- A video showing all the sites of resection in the full extent should be shot at the end of the MI-CRS phase and submitted to the regulatory committee for each patient.

**Mini-laparotomy phase**

At the end of the MI-CRS, the abdomen will be opened though a midline mini-laparotomy that extends from mid-way between the xiphoid process to the umbilicus to mid-way between the umbilicus and pubic symphysis. The following should be performed in all patients.

1. All the resected areas will be inspected for the presence of any residual disease. The 13- region residual PCI will be recorded in the PCI form that has provision for the same.
2. The whole colon and rectum will be palpated for the presence of nodules. The entire small bowel will be inspected for presence of tumor nodules. The lesser omentum will be inspected and palpated for presence of disease. If a splenectomy is not performed, then the splenic hilum and posterior edge will be both inspected for presence of disease. The Morrison’s pouch, gall bladder and foramen of Winslow will be inspected for presence of disease. Liver mobilization could be done if deemed necessary by the surgeon but is not mandatory. The ovaries if present will be inspected for presence of disease.
3. A total omentectomy preserving the gastroepiploic arch will be performed for all patients even if absence of gross disease if the same was not done during MI-CRS. If the arch is directly involved or resection deemed necessary by the surgeon, it will be resected. The falciform ligament and umbilical round ligament will be resected in all patients irrespective of the presence of disease if the same was not done during MI-CRS.
4. Any newly identified areas of disease will be resected performing the necessary peritonectomies and visceral resections. Conversion to full laparotomy should be done if any new disease is detected during MI-CRS.
5. The specimens will be sent for histopathological evaluation.
6. All port sites will be resected at the end of the procedure and sent for pathologic evaluation.

At the end of the procedure, HIPEC will be performed if indicated by the open or the closed method according to the preference of the surgeon.

The postoperative management will be according to the protocol at each institution.

**7.3.2 Histopathological evaluation**

Each resected peritoneal region should be evaluated separately. Two or more representative sections should be taken from each region. The report should provide the findings of each peritoneal region separately.

Ascitic fluid or peritoneal washings will undergo cytologic analysis and cell block analysis if required.

**7.3.3 Additional treatment(s)**

Additional systemic treatments will be at the discretion of the treating clinicians but will be captured in the CRF and submitted for each case.

**7.3.4 Follow-up/ surveillance**

Routine follow-up of all patients will be done. Depending on the primary tumor site, surveillance will involve physical examination, blood tests and cross-sectional imaging as deemed necessary by the treating clinician. The frequency will be every 3-6 months for the first two years and every 6-12 months for the next three years, depending on the primary tumor site.

##### **7.4** **Description of activities performed by sponsor representatives**

- SPSO will provide funds for administrative work to the principal investigators

##### **7.5. Any known or foreseeable factors that can compromise the outcome**

All the surgeons are experienced in performing CRS and minimally invasive surgery. This would obviate any unexpected injuries or adverse events.

##### **7.6.1. Address recommended follow-up for the subjects**

**Dr. Aditi Bhatt**

Sr consultant Surgical Oncologist,

Shalby Hospital, Ahmedabad

India-380015

#### **7.6.2 Monitoring plan**

The clinical investigation will be monitored by the designated body at each investigation site. This will be done before the clinical investigation begins, during the clinical investigation, and after the clinical investigation has been completed, so as to ensure that the clinical investigation is carried out according to the CIP and that data is collected, documented, and reported according to the applicable ethical and regulatory requirements. Monitoring is intended to ensure that the subject’s rights, safety, and well-being are met as well as data in the CRF are complete, correct, and consistent with the source data.

The videos of staging laparoscopy and MI-CRS will be reviewed by an independent review committee. Individual institution data monitoring and safety committee will be ensure that investigators conform to ethical clinical and research practices laid down by the GCP.

### **8. Statistical design and analysis**

#### **8.1. Analysis population**

In open surgery, the rate of missed metastases is considered to be zero. So any amount of missed metastases in this study should be unacceptable. For the primary end-point, no comparative group is required.

For the secondary end-points, patients who were excluded because of contraindications to CRS, or extensive adhesions or prior CRS by laparotomy and those that were excluded after staging laparoscopy, will provide the comparator group.

The primary and secondary end-points will be evaluated on the per-protocol population and modified intention-to-treat population.

#### **8.2. Descriptive statistics**

Continuous variables will be recorded as medians and interquartile range. Categorical variables will be presented as numbers and proportions. Survival end points will be calculated from the day of surgery to the event. The event for overall survival is death and that for progression-free or relapse-free survival is any recurrence. Patients not experiencing the event will be censored at their last follow up. Time-to-event data will be measured using the Kaplan-Meier method and follow-up durations using the reverse Kaplan-Meier method.

#### **8.3. The significance level and the power of primary endpoint and the overall statistical testing strategy**

The study involves a convenient sampling; therefore, the significance level and the power will be low and not applicable to this study.

#### **8.4. Sample size calculation and justification**

The crux of the study is peritoneal metastases missed during MI-CRS. Considering that the patients that will be eligible for this study will be those with low volume disease that is potentially curable, the margin of error should be minimum. An incomplete surgery is this biggest negative prognostic factor in patients undergoing CRS irrespective of the primary tumor site. Therefore, the incidence of missed metastases should be 0%. Considering that 5% or fewer patients could have missed metastases and 10% should be the maximum tolerable limit with 95% confidence intervals, 88 patients will have to be recruited in the study. Therefore, if 88 patients are assessed and <4 have missed metastases, there would be 95% certainty that not more than 10% have missed metastases. To account for conversion during the course of MI-CRS and other unexpected problems, 100 patients will be recruited.

#### **8.5. Expected drop-out rate, such as withdrawal, lost to follow-up, death**

The impact of subject withdrawal is unlikely to affect the primary endpoint. The dropout rate has been estimated to be less than 10-15%. This is the usual rate in our previous experiences and we do not expect anything different in this study.

#### **8.6. Eligibility**

Surgeons well versed in both minimally invasive surgery for abdominal malignancies and cytoreductive surgery for peritoneal malignancies will be invited to participate. The greater emphasis is on the experience with cytoreductive surgery than minimally invasive surgery. Participation is not restricted to Indian centers. All surgeons will take ethics committee approval at their respective centers as required. The regulatory committee will monitor the quality of the surgery based on the two films submitted for each patient on a 3-monthly basis.

#### **8.7. Pass/fail criteria to be applied to the results of the clinical investigation**

Not applicable to the current study

#### **8.8. Missing data**

All dropouts will be accounted for and reported and the reason for the drop-outs will be mentioned.

We expect dropouts to be less than 10-15% in the study. The drop-out rate should not affect the primary end-point which is the proportion of patients with pathologically proven peritoneal disease that was not identified/resected during MI-CRS

### **9. Data management and protection**

Subjects who participate in the clinical investigation are coded with a specific clinical investigation identification number. All subjects are registered in a subject identification list (subject enrolment and identification list) that connects the subject’s name and personal number with a clinical investigation identification number.

All data will be registered, managed, and stored in a manner that enables correct reporting, interpretation, and verification. All clinical data will be entered into the CRF by authorized study site personnel designated by the Investigator, as required by the protocol.

Data query handling and internal quality checks to identify data that appear inconsistent, incomplete, or inaccurate will be performed by principle investigators.

#### **9.1. Case Report Form and methods for data entry and collection**

All clinical data will be entered into the CRF. Authorized study site personnel designated by the Investigator will be entering information in the CRF, as required by the protocol.

Appropriate training will be completed with the Investigator and all authorized study site personnel, before the study initiation, and before any study data is entered into the CRF.

The investigator or authorized designated staff will record subject data in the CRFs in a precise and accurate manner. Abbreviations should not be used. All data should be entered in English. The CRFs should be completed daily as soon as possible after the assessments.

The Investigator is responsible for ensuring the accuracy, completeness, legibility, and timeliness of the data recorded in the CRFs and for signing the CRFs no later than at the end of the clinical investigation. The data should be recorded as soon as they are generated.

In the process of ensuring data completeness and accuracy, source data verification (SDV) will be performed by the PI.

All data processed during the trial will only be identified by study subject number, thereby ensuring that the study subject’s identity remains unknown to unauthorized personnel. The Investigator will arrange for the retention of the study subject identification, the code list, and CRF for at least 5 years after the completion of the trial. Patient files and other source data will be kept in accordance with standard procedures at the site. All information concerning the trial should be stored in a safe place inaccessible to unauthorized personnel.

The Investigator and designated site personnel will have read/write access to the CRF.

It is the Investigator´s responsibility to ensure completion and to review and approve all CRFs.

#### **9.2. Procedures to maintain and protect subject privacy**

The subject identification documents, the code list, and the CRF data after being signed by the Investigator will be stored at the Investigational site by the Investigator in a safe and secure place.

#### **9.3. Procedures for data retention**

The Investigator will arrange for the data retention of the study subject identification, the code list, and CRF at the investigational site.

#### **9.4. Specified retention period**

At least 5 years after the completion of the clinical trial.

#### **9.5. Other aspects of clinical quality assurance**

##### **9.5.1. Source data**

The Investigator will complete and maintain source documents for each study subject participating in the trial. Data that are recorded directly in the CRF and not in the patient’s medical records are considered source data.

It is the responsibility of the Investigator to record essential information regarding the study subject’s participation in the trial in the medical records:

• Trial code

• Study subject screening number and/or study subject number

• That informed consent for participating in the trial was obtained

• All visits pertaining to the study

• All AE/ SAE

The monitoring plan specifies the scope and details of source data verification.

#### **9.6. Archiving**

The PI and sponsor will maintain the essential clinical investigation documents in the investigation site files archive and sponsor files archive, respectively. The PI will archive all local investigation documentation for at least 5 years or as long as stipulated by the local institution.

#### **9.7. Data protection**

The study team will follow the General Data Protection Regulation (EU ordinance 2016/679, GDPR) and other relevant legislation before any data transfer takes place.

The content of the patient consent form shall comply with relevant integrity and data protection legislation. In the subject information and the informed consent form, the subject will be given complete information about how the collection, use, and publication of their clinical investigation data will take place. The subject information and the informed consent form will explain how clinical investigation data are stored to maintain confidentiality in accordance with national data legislation.

All information processed by the sponsor will be pseudonymized and identified with <<Study code/Study ID>>.

The informed consent form will also explain that for verification of the data, authorized representatives of the sponsor, as well as relevant authority, may require access to parts of medical records or study records that are relevant to the clinical investigation, including the subject’s medical history.

### **10. Amendments to the CIP**

Study-related documents such as the CIP, CRFs, ICF, and other subject information, or other clinical study documents will be amended as needed throughout the clinical study, and a justification statement will be included with each amended section of a document. Proposed amendments to the CIP will be agreed upon between the Sponsor and PI.

The amendments to the CIP and the subject’s Informed Consent will be notified to, or approved by, the EC and regulatory authorities. The version number and date of amendments will be documented.

The amendment will identify the changes made, the reason for the changes, and whether implementing the amendment is mandatory or optional.

### **11. Deviations from the CIP**

Investigator(s) are not allowed to deviate from the CIP except if it is for the protection of the subject´s rights, safety, or well-being under emergency circumstances.

All such deviations shall be documented with an explanation and reported to the sponsor, the EC as soon as possible.

Deviations will be reviewed by the sponsor and reported to the appropriate regulatory bodies as required.

The investigator and the entire study team shall adhere to the approved protocol by the ethics committee.

### **12. Statements of compliance**

#### **12. 1. Compliance with the investigational plan, good clinical practice, and regulations**

The clinical investigation will be conducted in accordance with the clinical investigation plan, the ethical principles of the Declaration of Helsinki, and current national and international regulations governing this clinical investigation. This is to ensure the safety and integrity of the subjects as well as the quality of the data collected.

The study will be registered at www.ctri.nic.in and www.clinaltrials.gov before starting the study.

#### **12.2. Ethical review of the clinical investigation**

The clinical investigation will not commence until written approval/favorable opinion from the EC and regulatory authority have been received.

The final version of the informed consent form and other information provided to subjects must be approved, or given a written positive opinion by the EC. The EC must be informed of any changes in the CIP in accordance with the current requirements.

The Investigator will not start to enroll subjects or request informed consent from any subject before obtaining EC and regulatory authority approvals.

Any additional requirements imposed by the EC or regulatory authority will be followed in the clinical investigation.

### **13. Informed consent process**

#### **13.1. General process for obtaining informed consent**

The PI shall ensure that the subject is given full and adequate oral and written information about the clinical investigation, its purpose, any risks and benefits as well as inclusion and exclusion criteria. Subjects must also be informed that they are free to discontinue their participation in the clinical investigation at any time by providing a reason. Subjects shall be allowed to ask questions and be allowed time to consider the provided information and participation in the clinical investigation. If the person chooses to participate, both the subject and the investigator shall sign the informed consent form. A copy of the subject information as well as a copy of the informed consent form shall be provided to the subject. The subject’s signed and dated informed consent must be obtained before performing any activity specific to the clinical investigation. The process shall be documented in the subject’s source documents and the signed informed consents shall be maintained with the essential documents. If new information becomes available that can significantly affect a subject's future health and medical care that information shall be provided to the affected subject in written form. If new information is added to the clinical investigation, the subject has the right to reconsider whether he/she will continue their participation.

#### **13.2. Informed consent for the subject unable to provide or in an emergency case**

A legal representative of the subject should provide the informed consent to subject’s participation in the clinical investigation.

#### **14. Adverse events**

Adverse Events (AE) will be collected for the duration of the investigation, starting from the subject’s inclusion date and until the last study visit, and recorded in the CRF on the adverse event form. It is the responsibility of the Investigator to ensure that all information is correct and that all staff involved in the investigation are familiar with the definitions and procedures of AE reporting.

#### **14.1. Definitions**

##### **14.1.1. Adverse event**

An Adverse Event (AE) is an untoward medical occurrence, unintended disease or injury, or any untoward clinical signs, including an abnormal laboratory finding, in subjects, users, or other persons, in the context of a clinical investigation, whether or not related to the investigational device.

This definition includes events that are anticipated as well as unanticipated events.

The reporting of adverse events will be according to the common terminology criteria for adverse events (CTCAE-v5) <https://ctep.cancer.gov/protocoldevelopment/electronic_applications/docs/ctcae_v5_quick_reference_5x7.pdf>

**15.1.3. Serious adverse events**

A Serious Adverse Event (SAE) is any AE that led to any of the following:

a) Death,

b) Grade 3-4 adverse events according to CTCAE criteria

#### **16.2. Recording and Reporting**

##### **16.2.1. Recording**

The principal investigator will record:

- all AEs
- all SAEs;
- any new finding in relation to any of the above-mentioned events.

##### **16.2.2. Reporting**

All adverse event will be recorded in the CRF

The following information will be recorded: description of the course of adverse event, date of onset, duration, severity of adverse event, diagnosis of adverse event, treatment of adverse event, and outcome.

#### **16.3. List of foreseeable adverse events**

- Infection
- Internal bleeding
- Damage to tissue/organ

#### **16.4.** **Emergency contact details for reporting serious SAE**

Dr. Aditi Bhatt

Sr. Consultant Surgical Oncologist

Shalby Hospital, Ahmedabad

#### **16.5. Data monitoring committee (DMC) or regulatory committee**

The PI shall establish a DMC prior to starting the clinical investigation. This committee will review the videos that are submitted at different stages of the surgical intervention and score the quality on a separate form **(Annexures 1 and 2)**.

The responsibilities of the DMC are:

- Assess the quality of the staging laparoscopy and MI-CRS videos using the designated forms
- Monitoring the interim and final study results

The DMC will have the following members:

| **Sl. No.** | **Member** |
| --- | --- |
| 1. | Prof. Ramakrishnan AS  Professor (GI & HPB oncology)  Dept. of Surgical Oncology  Cancer Institute (WIA)  Chennai, India |
| 2 | Dr. Prasanth Penumadu  Head of the Department, Surgical Oncology Sri Venkateswara Institute of Cancer Care and Advanced Research (SVICCAR),  A Unit of Tata Cancer Care Foundation,  Tirupati - 517501 |
| 3 | Dr. Ashvin Rangole  Clinical director and surgical oncologist,  CARE CHL hospitals,  Indore, Madhya Pradesh |

### **17. Vulnerable population**

Patients who do not meet the inclusion criteria will not be considered for this study.

### **18. Suspension or premature termination of the clinical investigation**

#### **18.1. Criteria and arrangements**

The investigator may also discontinue participation in the clinical study with appropriate written notice to the PI.

A PI, EC, or regulatory authority may suspend or prematurely terminate participation in a Clinical Investigation at the investigation sites for which they are responsible. If suspicion of an unacceptable risk, including serious health threat to subjects, arises during the clinical investigation, or when so instructed by the EC or regulatory authorities, the PI shall suspend the clinical investigation while the risk is assessed.

If suspension or premature termination occurs, the terminating party will justify its decision in writing and promptly inform the other parties with whom they are in direct communication.

The PI may suspend or prematurely terminate the entire clinical investigation for significant and documented reasons, such as when recommended by the DMC.

The EC may suspend or prematurely terminate the clinical investigation at the investigation site.

If suspicion of an unacceptable risk to subjects arises during the clinical investigation, or when so instructed by the EC, the PIs will suspend the clinical investigation while the risk is assessed. The PIs will terminate the clinical investigation if an unacceptable risk is confirmed. The PIs will inform all investigators.

If, in the opinion of the investigator, the clinical observations in the clinical investigation suggest that it may be unsafe to continue the investigation at the site, the investigator may terminate participation in the investigation after consultation with the PIs. A written statement fully documenting the reasons for such termination will be provided to the PIs. If the clinical investigation is prematurely terminated, the investigator shall promptly inform the subjects and take necessary steps to finalize their engagement in the clinical investigation. All relevant investigation material must be collected, and accountability completed.

If the clinical investigation is interrupted or terminated prematurely the PI shall report to the EC within 15 days together with a justification. A clinical investigation report will be prepared within three months of the early termination or temporary halt, irrespective of the results. If the clinical investigation is restarted within three months of the temporary halt, the PI does not have to submit a clinical investigation report until the clinical investigation has been completed.

The final clinical investigation report shall include detail concerning the temporary halt.

#### **18.2. Requirements for subject follow-up and continued care**

The hospital stay will depend on the clinical condition of each patient and other logistics. The follow-up of the subjects will be as per the protocol at each institution. The follow-up data will be provided by each investigator on a 6-monthly basis.

### **19. Publication policy**

The publication policy should cover authorship, acknowledgements, and review procedures for scientific publications. If there is a department or institution policy, or agreement, the protocol can refer to it.

The PI can publicize this study with the written consent of the sponsor.

### **20. List of personnel involved in the study and designations at the time of study**

| Title | Name | Designation | Role & Responsibility |
| --- | --- | --- | --- |
| Doctor | Dr. Aditi Bhatt  Sr. Consultant Surgical Oncologist  Shalby Hospital, Ahmedabad  | Principal Investigator | Conduct of the study adhering to good clinical practice, Management of the day-to-day conduct of the clinical investigation, ensure data integrity, rights, safety, and well-being of the subjects involved in the clinical investigation.  Coordinating with DMC and EC.  Informing all team members about new requirements. |
| Doctor | Dr. Sanket Mehta  Director, Specialty Surgical Oncology Hospital and Research Centre, Ghatkopar, Mumbai  | Principal Investigator | Management of the day-to-day conduct of the clinical investigation, ensure data integrity, rights, safety, and well-being of the subjects involved in the clinical investigation.  Coordinating with DMC and EC. |
| Doctor | **Dr. Mufaddal Kazi**  Associate Professor, Surgical Oncology  Department of Colorectal and gastrointestinal surgery,  Tata Memorial Hospital, Parel, Mumbai  | Co-Investigator | Patient enrolment,  Performance of the procedure sticking to EC approved plan, ensuring safety and well-being of the subjects.  Follow up.  Data collection and analysis. |
| Doctor | **Dr. Swapnil Patel**  Surgical Oncologist  Clinical Lead - GI, HPB & Peritoneal Surface Oncology  Medical Director, Upkar Cancer Institute (unit of Upkar Hospital Pvt Ltd) Varanasi  | Co-Investigator | Patient enrolment,  Performance of the procedure sticking to EC approved plan, ensuring safety and well-being of the subjects.  Follow up.  Data collection and analysis. |
| Doctor | **Dr. Deepti Mishra**  Consultant Surgical oncology,  Thangam cancer centre, Namakkal, Tamil Nadu  | Co-Investigator | Patient enrolment,  Performance of the procedure sticking to EC approved plan, ensuring safety and well-being of the subjects.  Follow up.  Data collection and analysis. |
| Doctor | **Dr. Dileep Damodaran**  Consultant, Surgical Oncology,  MVR cancer centre and research institute,  Calicut, India  | Co-Investigator | Patient enrolment,  Performance of the procedure sticking to EC approved plan, ensuring safety and well-being of the subjects.  Follow up.  Data collection and analysis. |
| Doctor | Prof. Ramakrishnan AS  Professor (GI & HPB oncology)  Dept. of Surgical Oncology  Cancer Institute (WIA)  Chennai, India | DMC/ regulatory committee | Quality assessment  Periodic review of the results |
| Doctor | Dr. Prasanth Penumadu  Head of the Department, Surgical Oncology Sri Venkateswara Institute of Cancer Care and Advanced Research (SVICCAR),  A Unit of Tata Cancer Care Foundation, Tirupati - 517501 | DMC/ regulatory committee | Quality assessment  Periodic review of the results |
| Doctor | Dr. Ashvin Rangole  Clinical director and surgical oncologist,  CARE CHL hospitals,  Indore, Madhya Pradesh | DMC/ regulatory committee | Quality assessment  Periodic review of the results |

### **21. Conclusion**

The study will be conducted as per guidelines as per the Declaration of Helsinki and the principle of Good clinical practice.

Investigators will follow the procedure at each stage of the study, namely, taking consent of the subject, recruiting only those subjects who fulfil all the criteria, proper filling of CRFs, maintaining investigational reports of all subjects, maintaining investigational products, giving proper instructions to the subjects during study, etc.

### **22. Annexure**

A Form for evaluating the quality of staging laparoscopy

B Form for evaluating the quality of MI-CRS

### **23. Signature page**

I have read this protocol and confirm that to the best of my knowledge, it accurately describes the conduct of the study. I adhere to the protocol for the conduct of the clinical investigation.

**____________________	___________________	__________________**

**Name	Signature	Date**

**Principal Investigator**

**Aditi Bhatt**
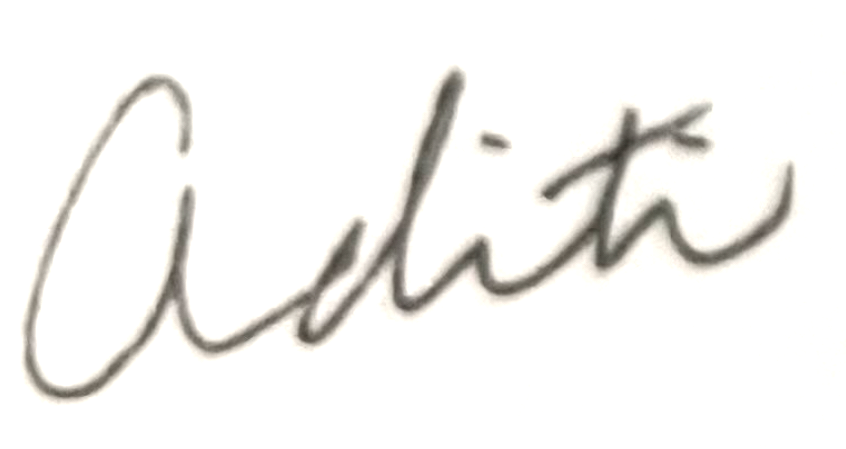
**5/9/2024**

**Name	Signature	Date**

**Principal Investigator:**

**Sanket Mehta**
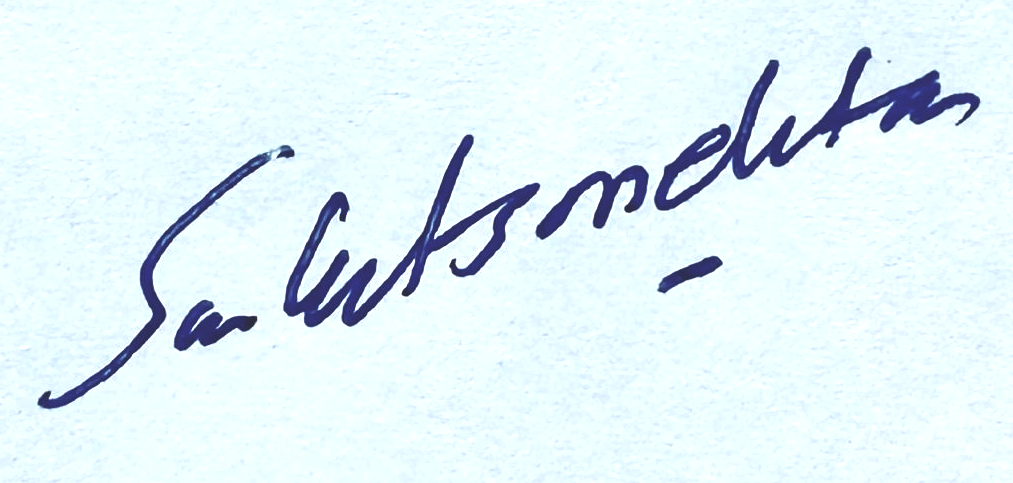
**5/9/2024**

**Name	Signature	Date**

**Co-Investigator:**

**Mufaddal Kazi**
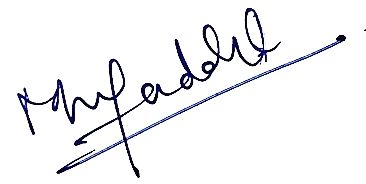
**5/9/2024**

**Name	Signature	Date**

7. Pedra Nobre S, Mueller JJ, Gardner GJ*, et al*. Comparison of minimally invasive versus open surgery in the treatment of endometrial carcinosarcoma

*International Journal of Gynecologic Cancer* 2020;**30:**1162-1168.

8. Arjona-Sanchez, A, Esquivel, J, Glehen, O, Passot, G, Turaga, KK, Labow, D, et al.. A minimally invasive approach for peritonectomy procedures and hyperthermic intraperitoneal chemotherapy (HIPEC) in limited peritoneal carcinomatosis: the American Society of Peritoneal Surface Malignancies (ASPSM) multi-institution analysis. Surg Endosc 2019;33:854–60. <https://doi.org/10.1007/s00464-018-6352-4>.

9. Passot G, Bakrin N, Isaac S, Decullier E, Gilly FN, Glehen O, Cotte E. Postoperative outcomes of laparoscopic vs open cytoreductive surgery plus hyperthermic intraperitoneal chemotherapy for treatment of peritoneal surface malignancies. Eur J Surg Oncol. 2014 Aug;40(8):957-62. doi: 10.1016/j.ejso.2013.10.002. Epub 2013 Oct 16. PMID: 24209429.

10. Arjona-Sanchez A, Aziz O, Passot G, Salti G, Esquivel J, Van der Speeten K, Piso P, et al. Laparoscopic cytoreductive surgery and hyperthermic intraperitoneal chemotherapy for limited peritoneal metastasis. The PSOGI international collaborative registry. Eur J Surg Oncol. 2021 Jun;47(6):1420-1426. doi: 10.1016/j.ejso.2020.11.140. Epub 2020 Dec 2. PMID: 33298341.

11. Durán-Martínez, M., Gómez-Dueñas, G., Rodriguez-Ortíz, L. *et al.* Laparoscopic versus open approach for interval cytoreductive surgery and hyperthermic intraperitoneal chemotherapy (HIPEC) in advanced epithelial ovarian cancer: a matched comparative study. *Surg Endosc* **38**, 66–74 (2024). <https://doi.org/10.1007/s00464-023-10508-w>

12.Arjona-Sanchez A, Aziz O, Passot G, Salti G, Serrano A, Esquivel J, Van der Speeten K, et al. Laparoscopic cytoreductive surgery and hyperthermic intraperitoneal chemotherapy: Long term oncologic outcomes from the international PSOGI registry. Eur J Surg Oncol. 2023 Oct;49(10):107001.

13. Nitecki R, Rauh-Hain JA, Melamed A, Scambia G, Pareja R, Coleman RL, Ramirez PT, Fagotti A. Laparoscopic cytoreduction After Neoadjuvant ChEmotherapy (LANCE). Int J Gynecol Cancer. 2020 Sep;30(9):1450-1454. doi: 10.1136/ijgc-2020-001584. Epub 2020 Jul 20. PMID: 32690591; PMCID: PMC7493891.

14. Uwins C, Assalaarachchi H, Bennett K, Read J, Tailor A, Crawshaw J, Chatterjee J, Ellis P, Skene SS, Michael A, Butler-Manuel S. MIRRORS: a prospective cohort study assessing the feasibility of robotic interval debulking surgery for advanced-stage ovarian cancer. Int J Gynecol Cancer. 2024 Jun 3;34(6):886-897. doi: 10.1136/ijgc-2024-005265. PMID: 38561194.

15. Gueli Alletti S, Bottoni C, Fanfani F, et al. Minimally invasive interval debulking surgery in ovarian neoplasm (MISSION trial-NCT02324595): a feasibility study. Am J Obstet Gynecol 2016;214:503.e1–503.e6.

16.Jacquet P, Sugarbaker PH. Clinical research methodologies in diagnosis and staging of patients with peritoneal carcinomatosis. Cancer Treat Res. 1996;82:359-74. doi: 10.1007/978-1-4613-1247-5_23. PMID: 8849962

17. Bhatt A, Yonemura Y, Benzerdjeb N, Mehta S, Mishra S, Parikh L, Kammar P, Shah MY, Prabhu A, Shaikh S, Patel MD, Isaac S, Glehen O. Pathological assessment of cytoreductive surgery specimens and its unexplored prognostic potential-a prospective multi-centric study. Eur J Surg Oncol. 2019 Dec;45(12):2398-2404. doi: 10.1016/j.ejso.2019.07.019. Epub 2019 Jul 16. PMID: 31337527.

18.Finch L, Chi DS. An overview of the current debate between using minimally invasive surgery versus laparotomy for interval cytoreductive surgery in epithelial ovarian cancer. J Gynecol Oncol. 2023 Sep;34(5):e84.

**Annexure 1**

Quality assessment of staging laparoscopy

Reviewer name: ______________________

Date of Staging laparoscopy: ______________________

| **Basic background information to be provided to the reviewer** |
| --- |
| Name/identifier |
| Centre |
| Country |
| Age |
| Sex |
| Primary tumour site |
| For colorectal cancer-exact site of the primary |
| For ovarian cancer- exact site (ovaries, FT, peritoneum) |
| Timing of peritoneal malignancy (synchronous/metachronous) |
| Date of diagnosis of the primary |
| Date of diagnosis of peritoneal malignancy |
| TNM stage at diagnosis |
| FIGO stage at diagnosis (for ovarian cancer) |
| Prior CRS |
| Date of prior CRS |
| Prior CRS+HIPEC |
| Date of prior CRS+HIPEC |
| Prior surgery (not CRS) |
| Prior surgical score |
| Previous chemotherapy |
| Number of lines of previous chemotherapy |

Film quality

| Clarity and resolution | Acceptable |
| --- | --- |
|  | Below par |
|  | Not acceptable |
| If rated ‘not acceptable’, state reasons |  |
| Any other general comments |  |

Staging laparoscopy

| PCI region | Visualized clearly | All boundaries seen | Disease seen/not seen | Concurrence with PCI score | Critical structure | Visualized/ not visualized |
| --- | --- | --- | --- | --- | --- | --- |
| 0 |  |  |  |  | Transverse colon |  |
| 1 |  |  |  |  | Morrison’s pouch |  |
| 2 |  |  |  |  | Porta hepatis and attachment of lesser omentum to the liver |  |
| 3 |  |  |  |  | Splenic hilum and posterior edge of the spleen |  |
| 4 |  |  |  |  | Left paracolic gutter |  |
| 5 |  |  |  |  | Left iliac fossa and sigmoid mesentery |  |
| 6 |  |  |  |  | Pouch of Douglas in males and females, vesicovaginal space in females |  |
| 7 |  |  |  |  | Right iliac fossa and appendix |  |
| 8 |  |  |  |  | Right paracolic gutter |  |
| 9 |  |  |  |  | Root of the mesentery |  |
| 10 |  |  |  |  | Root of the mesentery |  |
| 11 |  |  |  |  | Root of the mesentery |  |
| 12 |  |  |  |  | Root of the mesentery |  |

**Annexure 2**

Quality assessment of MI-CRS

Reviewer name: ______________________

Date of MI-CRS: ______________________

| **Basic background information to be provided to the reviewer** |
| --- |
| Name/identifier |
| Centre |
| Country |
| Age |
| Sex |
| Primary tumour site |
| For colorectal cancer-exact site of the primary |
| For ovarian cancer- exact site (ovaries, FT, peritoneum) |
| Timing of peritoneal malignancy (synchronous/metachronous) |
| Date of diagnosis of the primary |
| Date of diagnosis of peritoneal malignancy |
| TNM stage at diagnosis |
| FIGO stage at diagnosis (for ovarian cancer) |
| Prior CRS |
| Date of prior CRS |
| Prior CRS+HIPEC |
| Date of prior CRS+HIPEC |
| Prior surgery (not CRS) |
| Prior surgical score |
| Previous chemotherapy |
| Number of lines of previous chemotherapy |

Film quality

| Clarity and resolution | Acceptable |
| --- | --- |
|  | Below par |
|  | Not acceptable |
| If rated ‘not acceptable’, state reasons |  |
| Any other general comments |  |

MI-CRS

| PCI region | Resected/not resected | Resection of region partial/ complete | Residual disease (seen/not seen) | Residual disease score | Critical structure | Visualized/ not visualized |
| --- | --- | --- | --- | --- | --- | --- |
| 0 |  |  |  |  | Transverse colon |  |
| 1 |  |  |  |  | Morrison’s pouch |  |
| 2 |  |  |  |  | Porta hepatis and attachment of lesser omentum to the liver |  |
| 3 |  |  |  |  | Splenic hilum and posterior edge of the spleen |  |
| 4 |  |  |  |  | Left paracolic gutter |  |
| 5 |  |  |  |  | Left iliac fossa and sigmoid mesentery |  |
| 6 |  |  |  |  | Pouch of Douglas in males and females, vesicovaginal space in females |  |
| 7 |  |  |  |  | Right iliac fossa and appendix |  |
| 8 |  |  |  |  | Right paracolic gutter |  |
| 9 |  |  |  |  | Root of the mesentery |  |
| 10 |  |  |  |  | Root of the mesentery |  |
| 11 |  |  |  |  | Root of the mesentery |  |
| 12 |  |  |  |  | Root of the mesentery |  |
